# Evaluation of international guidance for the community treatment of complex emotional needs: A systematic review

**DOI:** 10.1101/2022.02.14.22270639

**Authors:** Nicholas Wong Zhan Yuen, Phoebe Barnett, Luke Sheridan Rains, Sonia Johnson, Jo Billings

## Abstract

**Background:** Guidelines for the treatment and management of “personality disorder” were introduced to provide guidance on best practice from evidence and views of key stakeholders. However, this guidance varies as there is yet to be an overall, internationally recognised consensus on the best mental health care for people with complex emotional needs (CEN - our preferred working term for the needs of people using services for or related to “personality disorder”).

**Aims:** We aimed to identify and synthesise recommendations made by different mental health organisations from across the world on community treatment for people with CEN.

**Methods:** This systematic review consisted of three stages: 1. systematic literature and guideline search, 2. quality appraisal, and 3. data synthesis. We combined a search strategy involving both systematic searching of bibliographic databases and supplementary search methods of grey literature. Key informants were also contacted to further identify relevant guidelines. Codebook thematic analysis was then conducted. The quality of all included guidelines was assessed and considered alongside results.

**Results:** After synthesising 29 guidelines from 11 countries and 1 international organisation, we identified four main domains, with a total of 27 themes. Important key principles on which there was consensus included continuity of care, equity of access, accessibility of services, availability of specialist care, taking a whole systems approach, trauma informed approaches, and collaborative care planning and decision making.

**Conclusions:** Existing international guidelines shared consensus on a set of principles for the community treatment of CEN. However, half of the guidelines were of lower methodological quality, with many recommendations not backed by evidence.

## Introduction

“Personality disorder” is described in the fifth edition of the diagnostic statistical manual of mental disorders (DSM-5) [1] as “an enduring pattern of inner experience and behaviour that deviates markedly from the expectations of the individual’s culture” (p. 645). Individuals with a diagnosis of “personality disorder” have been estimated to have shorter lifespans compared to the general population [2, 3]. In 2009, the World Health Organisation (WHO) estimated the prevalence of “personality disorder” as 6.1% [4], however, only thirteen countries were included in its survey with six being from western Europe. More recently, Winsper and colleagues [5] reviewed data from 21 different countries and estimated that “personality disorder” had a global pooled variance of 7.8%. In the United Kingdom (UK), it has been estimated that two in every five patients presenting in secondary care services might meet diagnostic criteria of “personality disorder” [6].

The diagnosis of “personality disorder” is controversial, with multiple service user commentaries expressing the term as pejorative, associated with negative staff attitudes and therapeutic hopelessness. While some have acknowledged that receiving a diagnosis was a ‘turning point’ where they were able to conceptualise and validate their experiences [7], the label is still often seen by service users as stigmatising and associated with exclusion from services and a lack of effective care [7, 8]. Hence, in this paper we have followed other recent literature in choosing to use an alternative working term – Complex Emotional Needs (CEN) instead of “personality disorder” to describe the range of difficulties experienced by people who may use services for “personality disorder” or receive this diagnosis: we advocate future research to develop acceptable and accurate ways of characterising these difficulties for research and practice. Where other research and clinical guidance has explicitly used the term “Personality Disorder” we have retained the use of this term for clarity.

Lack of a broad and high-quality evidence base is a significant contributor to lack of standardisation in treatment and management of CEN. Research has mainly focused on people with a diagnosis of “borderline” or “antisocial personality disorder” [9]. The primary focus in research on community-delivered treatment has been on psychological interventions, such as cognitive behavioural therapy and dialectical behaviour therapy [10]. A recent scoping review by Ledden et al. [11] found that various forms of psychological therapy targeting CEN appear to be of equivalent effectiveness in leading to significant reductions in symptoms and self-harm. However, such therapies are often time-limited and narrow in their inclusion criteria, and there is a lack of evidence on how best to support people with CEN in the long-term, or on interventions focused on social needs. Little high-quality research has focused on treating people with comorbidities such as psychosis or substance-misuse, on trauma-focused interventions for this group, or on parents with CEN and younger or older people. Research on peer support and on interventions that are co-produced with people with relevant lived experience is also lacking. A wide variety of medications have also been used to treat CEN (anticonvulsants, dopaminergics, anti-psychotics, antidepressants, and even omega-3 fatty acid) [9,12]. However, pharmacological therapies are more contentious with lesser evidence to back their efficacy. A recent systematic review conducted by Hancock-Johnson *et al*. [12] looked at the treatment of borderline CEN using pharmacotherapies and concluded that there was insufficient high-quality evidence for an evidence-based decision about the use of medication for CEN to be made. The authors attributed these inconclusive results to the poor methodological rigour and lack of transparency of the reviewed studies.

In the UK in 2003 a key policy implementation document was published by the National Institute for Mental Health England (NIMHE) entitled “Personality Disorder: No Longer A Diagnosis of Exclusion” [13]. This publication concluded that there was a severe lack of dedicated services for individuals with CEN and established a policy programme aimed at developing services for people with CEN on a basis of equity with other mental health conditions. Guidance included the establishment of specialist community services and provision of additional training programmes for staff in both specialist and generic services. This has led to the development of dedicated services for individuals with CEN with a fivefold increase in such services observed in the UK between 2002 and 2015 [14]. However, recent studies by Trevillion et al. [15] and Foye et al. [16] where 30 service users and 50 clinicians respectively were interviewed on the needs of service users and clinicians’ perspectives of best practice community care for individuals with CEN concluded that continuity of care, stigmatising treatment, especially in generic services, lack of access to specialist care and lack of a holistic and personalised focus are among the major persisting problems in care for this group, with previous guidance implemented to quite a limited extent.

In Europe, the Mental Health Declaration for Europe was a testament of the WHO European Region acknowledging the importance of mental health [17]. This declaration urged WHO member states to give higher priority to mental health issues by introducing policies that raise awareness of mental health, collectively tackling stigma and discriminatory practices, and improving existing mental health care systems [17]. This was one of the foundations for the establishment of the European Society for the Study of Personality Disorders (ESSPD) in 2010, an international coalition formed to develop the limited evidence base on care for CEN within Europe and to promote dissemination of evidence-based treatment services [18]. These initiatives reflect an increased worldwide interest in endeavours to treat CEN more effectively and equitably, which has led to some countries publishing some form of guidance on the management and treatment of CEN. However, these different international guidelines have not been compared or their quality systematically assessed.

Clinical practice guidelines have been defined by the United States Institute of Medicine as “systematically developed statements to assist practitioners and patients in choosing appropriate health care for specific clinical conditions” [19]. Clinical practice guidelines are intended to ensure that the quality and content of services offered are standardised to a certain degree, allowing patients to have access to the same standard of care from the health organisation [20, 21]. However, the method for developing such guidelines tends to vary among organisations. Four types of practice guideline have been listed by Woolf [22]: informal consensus guideline, formal consensus guideline, evidence-based guideline, and explicit guideline. Each has a different method of development, with evidence-based guidelines and explicit guidelines tending to be of a higher quality than informal consensus and formal consensus guidelines. Evidence-based and explicit guidelines present recommendations based on scientific evidence and not solely on experts’ opinion.

The ESSPD conducted an initial review on CEN guidelines published within Europe including nine guidelines from eight countries. This review was the first of its kind, providing an overview of practices adopted by different countries in managing and treating CEN and discussing methods for developing a more rigorous guideline. The research gaps regarding management of CEN are accompanied by a lack of dissemination and implementation in routine clinical settings of the evidence that is available: guidelines have a significant potential role in promoting this [24].

The aim of this systematic review was to identify recommendations made by different organisations across the world on community treatment for people with CEN. The three main objectives of this review are as follows:

- To identify common recommendations made by the different guidance.
- To explore differences in recommendations made by the different guidance.
- To discuss how these findings could contribute to development of future clinical practice guidelines on managing CEN.

## Methods

This review was registered on PROSPERO, reference number: CRD42019143410.

### Study Design

Systematic reviews of evidence not only answer ‘what works’, but also guide the development of more evidence-informed policy [25]. By systematically reviewing guidelines, similar and different recommendations made within guidance can be identified, quality of evidence can be considered, and conclusions can be drawn having minimized biases [26].

### Search Strategy

We combined a search strategy involving both systematic searching of bibliographic databases and supplementary search methods of grey literature.

This review was initially part of a wider programme of research registered on PROSPERO (reference number: CRD42019131834) in which a number of reviews examining complex emotional needs were conducted [27, 28, 29]. A single search strategy was initially used for the whole programme, with the original searches conducted in March 2019. Additional specific searches for international guidance were then conducted in April 2021. A combination of MeSH and free text terms were used in this search (see Supporting Information S1 for the complete search strategy used in each bibliographic database and an example strategy of the original search). The search terms were developed with input from an experienced information scientist and no language limit was imposed. Seven databases were searched from 2003 up to April 2021: MEDLINE; Embase; PsycINFO; Cumulative Index to Nursing and Allied Health Literature (CINAHL); Social Policy and Practice; Health Management Information Consortium (HMIC); Applied Social Sciences Index and Abstracts (ASSIA).

We also searched fifteen guideline databases and mental health organisations that are primarily involved in CEN to look for consensus statements or clinical practice guidelines, or links to guidelines (see Supporting Information S2 for the full list of guideline databases and organisations included in this search). We followed principles for web-based searching as described by Briscoe [30]. Our searches were conducted on Google advanced search and the metasearch engine Dogpile to identify unpublished guidelines or consensus statements using a combination of the search terms ‘personality disorder’ and ‘guidelines’. Searches were screened up to a depth of ten pages each.

As international guidelines were of interest to this study, we selected eight additional languages for our web-based search, based on our team and experts’ advice about countries which have policies about CEN and would therefore be likely to have some published guidance. Using Google advanced search, we searched for relevant guidelines in French, German, Italian, Spanish, Danish, Norwegian, Portuguese, and Greek using Google translated search terms that were identical to the searches in English (see Supporting Information S3 for search terms used in different languages), with each language searched to a depth of four pages. Translated search terms were translated and subsequently back translated again to English to ensure that the terms carried the same meaning. Identified guidelines that were not in English were translated into English using the Google translate function on Google Chrome.

We also contacted key informants via email to further identify potentially relevant clinical practice guidelines or consensus statements that might not have been made available to the public or are less well known to non-native researchers. To identify key informants from Europe, North America, Australia, and New Zealand, we searched Google using the search terms ‘personality disorder’, ‘hospital/clinics’, and ‘treatment’. Within the retrieved webpage, we identified mental health professionals who specialise in the treatment of CEN. We prioritised contacting key informants who held a dual role of being a researcher in academic institutions and also a practitioner. We subsequently contacted a total of forty-seven key informants across eighteen countries over a period of two months (March – April, 2021). After the first email, a follow up email was sent two weeks later to key informants who had not responded, this process was repeated with a total of three emails being sent.

### Study Selection

Potential literature retrieved during the bibliographic searches were collated in Endnote X9 [31] and duplicates were removed. Two reviewers (NW and PB) independently screened all the titles and abstracts of the literature and selected potentially relevant articles. The selected articles were subsequently screened at full text by the two reviewers, with discrepancies resolved through discussion until consensus was reached. Potential guidelines and consensus statements retrieved from grey literature searches and key informants were collated in an excel spread sheet with duplicates manually removed. Initial screening of web searches was done by one reviewer (NW) with potentially relevant literature extracted into an excel-based form. Two reviewers (NW and LSR) then screened the full text of selected literature with discrepancies resolved by discussion with a third and fourth reviewer (PB and JB). We selected relevant literature based on meeting all of the following inclusion criteria:

a. Publication date of the literature after 2003, since NHS England only published and implemented ‘Personality Disorder: No Longer a Diagnosis of Exclusion’ in 2003, signifying a change in UK policy regarding the development of services for individuals with CEN. Literature published prior to 2003 was deemed to be less relevant to the needs of this study as we wanted to focus on current service contexts and practice, ensuring that guidelines are of contemporary relevance.
b. Guidelines that fulfilled the definition of a clinical practice guidelines according to the Institute of Medicine [19] or consensus statement made by organisations that are formally registered in the country of origin and are concerned with the treatment and management of CEN.
c. Guidelines or consensus statements that are intended to guide the treatment of individuals with a diagnosis of ‘personality disorder’ or who are experiencing symptoms related to this presentation.
d. Guidelines or consensus statements which relate to the provision, organisation, and delivery of services or treatment for CEN in the community.

### Data Extraction and Quality Appraisal

Key characteristics of the eligible literature were extracted by a reviewer (NW) using an excel-based form and the data was subsequently discussed and agreed upon with two other reviewers (PB and JB). We (NW and PB) conducted a quality appraisal of the selected literature using an amended version of the Appraisal of Guidelines for Research and Evaluation II framework (AGREE-II) [32]. Amendments were made based on the appropriateness of the question to the studies aims and are highlighted in the Supporting Information S4.

Appraisal of guidelines was first performed by an independent reviewer (NW). Upon completion, a third of the guidelines were randomly selected to be reviewed by the second reviewer (PB) and all disagreements were subsequently resolved through discussion between the two reviewers. The remaining two thirds of the literature was then re-evaluated according to the consensus made earlier. Individual domain scores for each guideline as well as an overall score were calculated. A unanimous decision was made by the research team for guidelines scoring 70% or more to be considered as high quality.

### Synthesis

Data were analysed and synthesised using thematic analysis, due to its flexibility and established guidance. Specifically, we used codebook thematic analysis [33] which combines both inductive and deductive approaches to the analysis of qualitative data. This allowed us to identify themes inductively from the data (rather than having a priori determined categories) but then to code all the data into the established coding framework in order to provide reliable frequency counts of how commonly certain themes were represented in the guidelines.

All forms of coding were facilitated by the latest version of QSR NVivo [34]. Six of the identified guidelines were initially coded line-by-line by two independent reviewers (PB and JB) who subsequently derived an initial list of potential codes. The potential codes were then organised into a provisional coding frame. The coding frame was reviewed and revised by a third reviewer (NW) in discussion with the first two reviewers, through line-by-line coding of the remaining guidelines. This iterative process was repeated with guidelines re-visited to ensure coding was performed sufficiently and appropriately whenever amendments were made to the coding frame. The summary of findings was then discussed between all three reviewers (NW, PB, and JB).

## Results

A total of 202 guidelines were identified from the bibliographic databases, from which 37 were duplicates. After screening the remaining 165 guidelines at title and abstract level, thirteen studies were retrieved for full text review with only one meeting the inclusion criteria.

We identified 94 guidelines initially from web-searches and recommendations from key informants. After manually removing 41 duplicates, the remaining 53 guidelines were retrieved for full text review. One guideline was unretrievable due to subscription issues. Twenty-eight of the remaining guidelines met the inclusion criteria. Out of the 24 excluded documents, half did not meet the criteria of being a clinical practice guideline or consensus statement, Figure 1 shows the PRISMA diagram of the review process [35].

**Figure 1.**
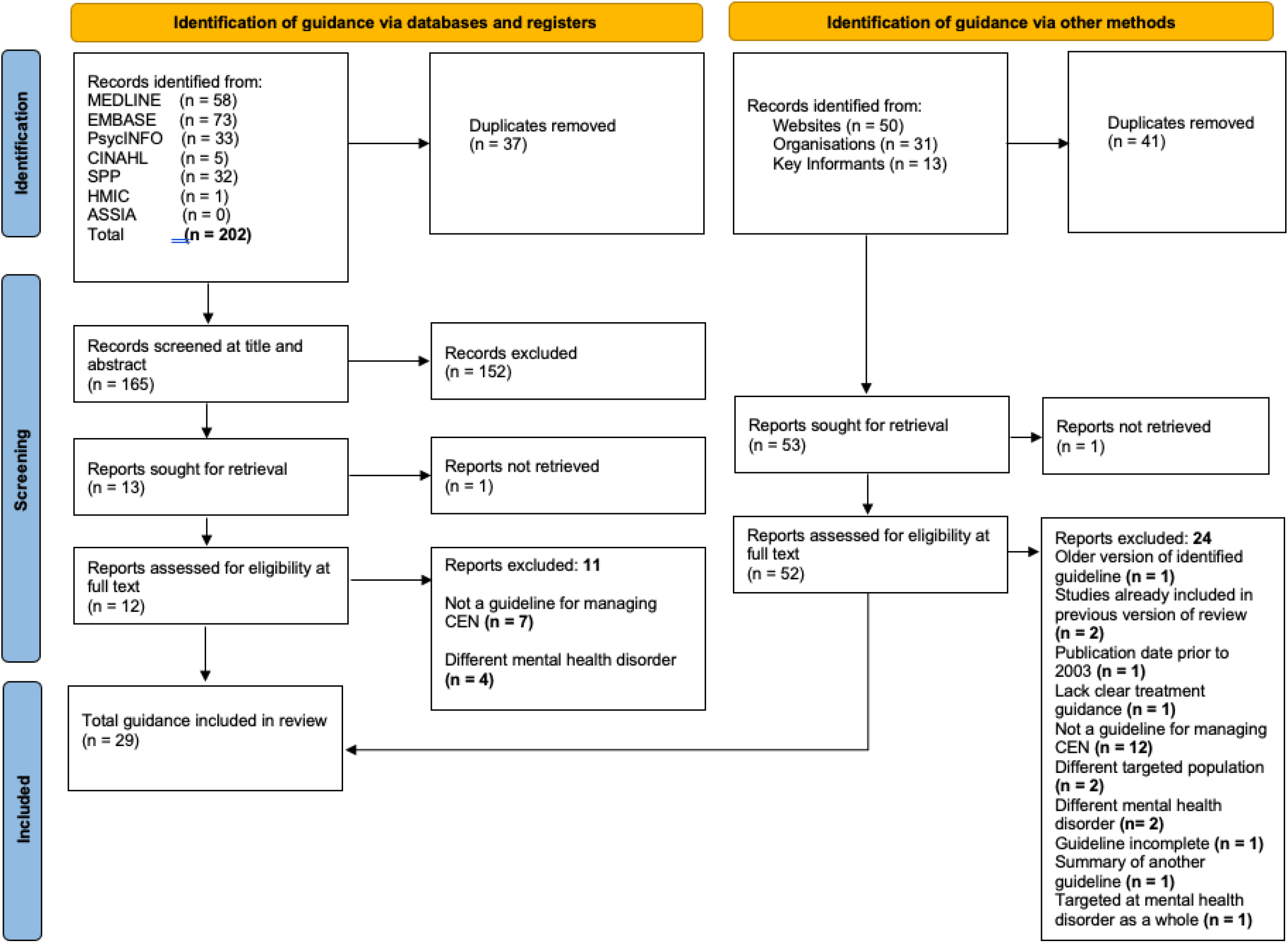
PRISMA diagram.

We identified in total 29 eligible guidelines and consensus statements. Consensus statements are comprehensive summary of opinions which are agreed upon by a panellist of subject matter experts [36], these statements might not necessarily be backed by evidence. Similar to guidelines, both types of documents’ primary goal is to provide readers with a form of guidance. Guidelines mentioned from here onwards consist of both guidelines and consensus statements.

### Characteristics of Included Guidelines

Twenty guidelines addressed “personality disorder” in general, eight focused on “borderline personality disorder” and one focused on “antisocial personality disorder”. Guidelines were from the UK (n = 10), Australia (n = 5), the Netherlands (n = 3), Spain (n = 3), with the remaining guidelines (n = 1) from USA, Denmark, Finland, Switzerland, Italy, Canada, Sweden, and an international organisation. Most of the guidelines are produced by governmental bodies (n = 24) with the remaining from voluntary sectors (n = 5). A summary of the characteristics of included guidelines can be found in Table 1.

**Table 1.**
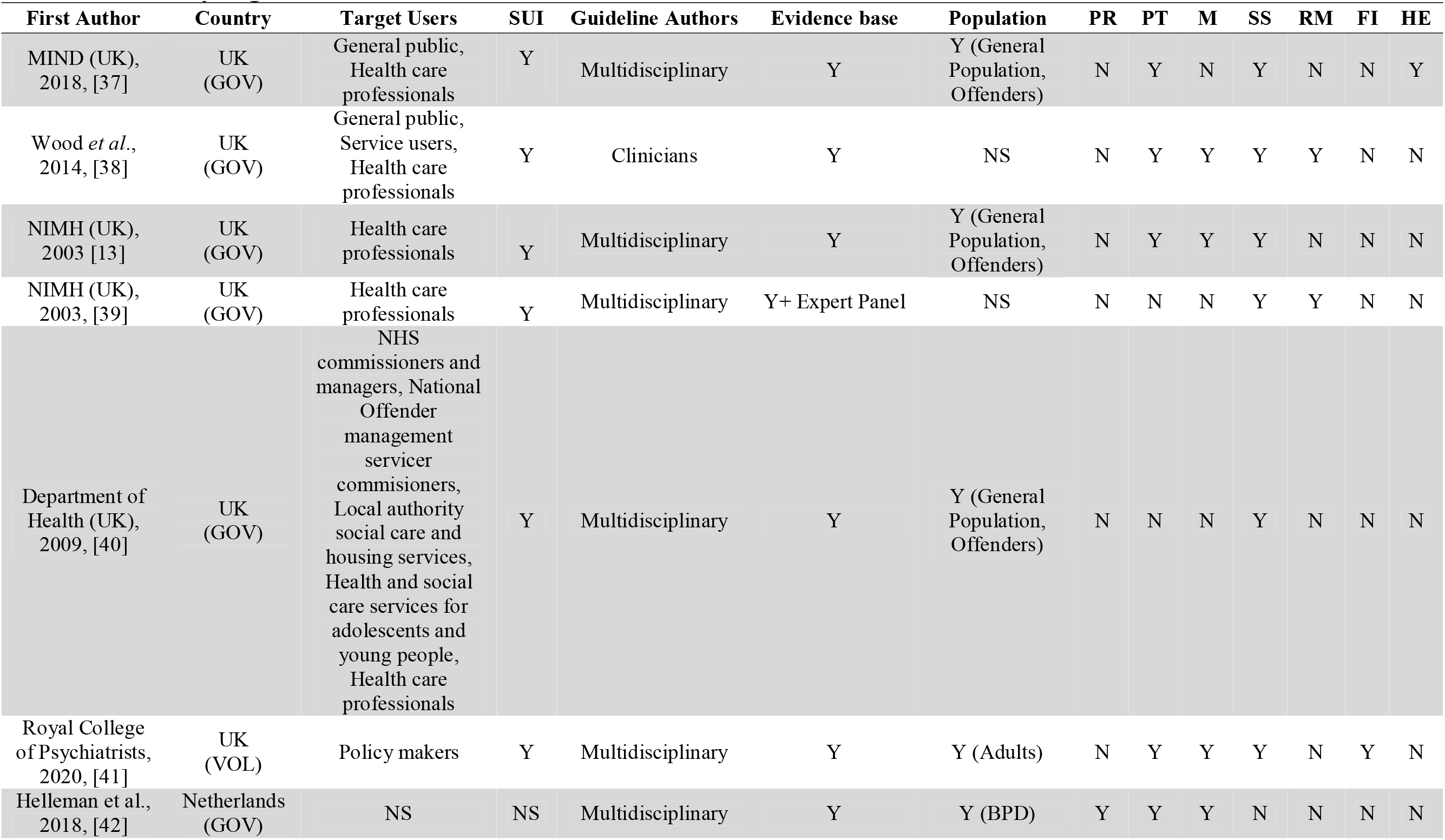
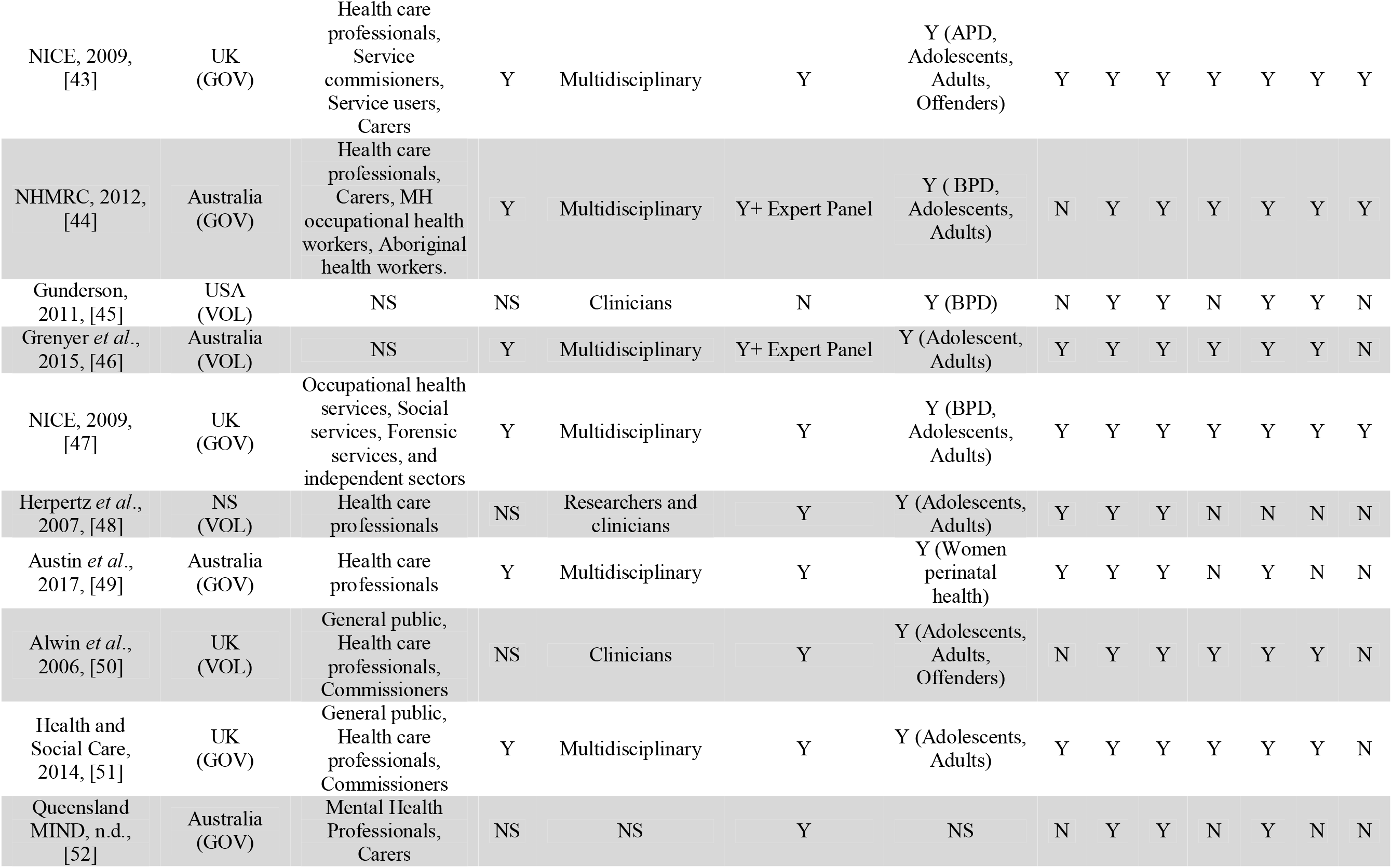
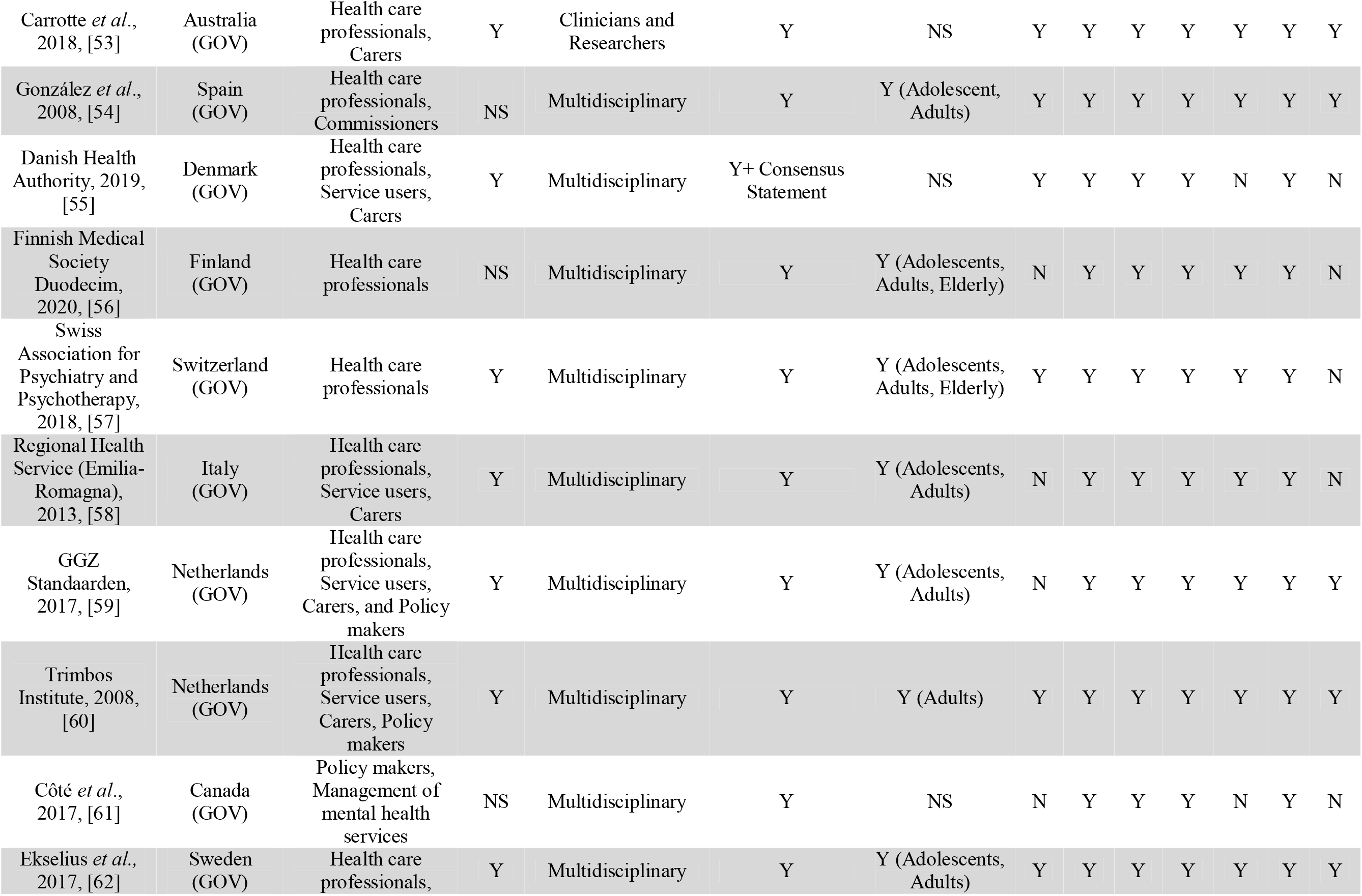
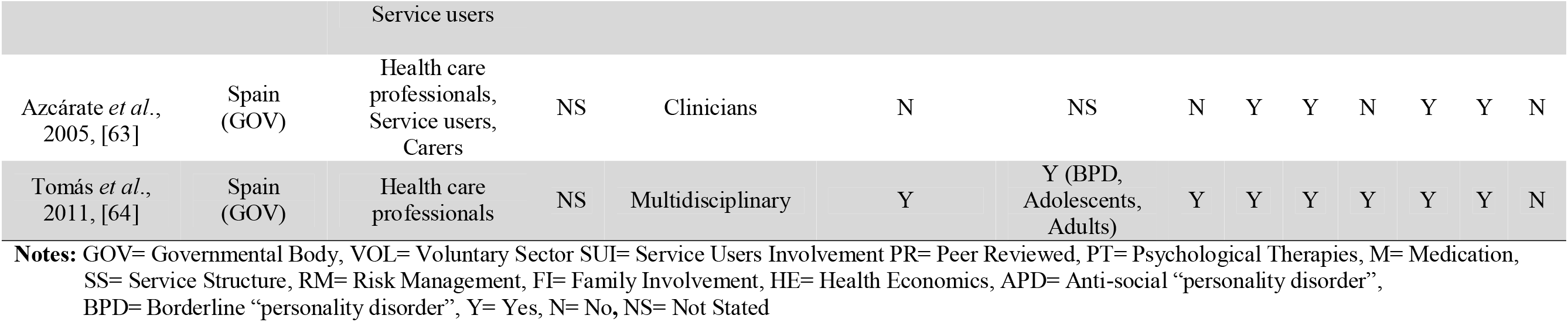
Summary of guideline’s characteristics.

### Quality Appraisal

Twelve of the 29 eligible guidelines were assessed to be of high quality. Individual domain scores of each guideline can be found in Table 2.

**Table 2.**
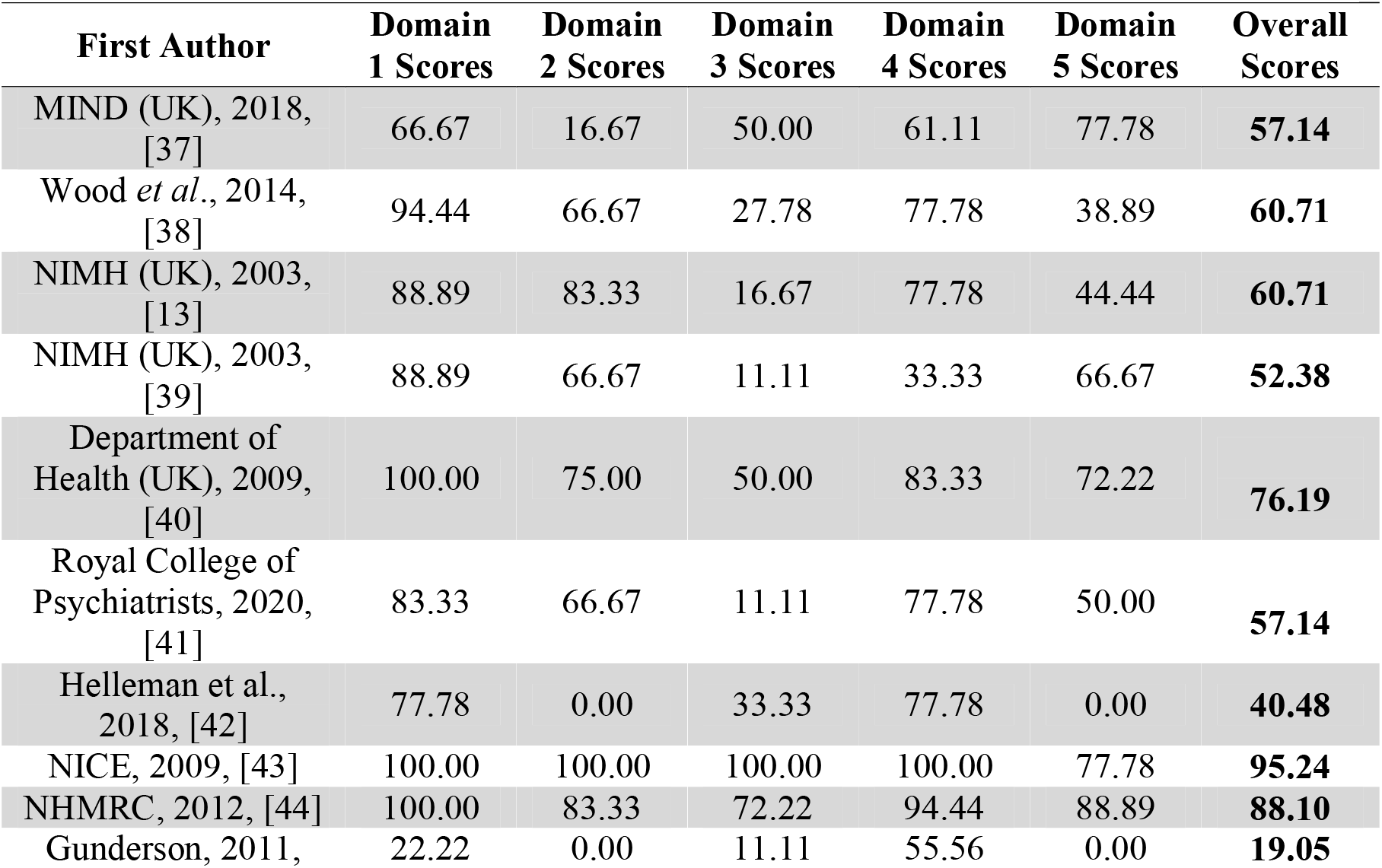
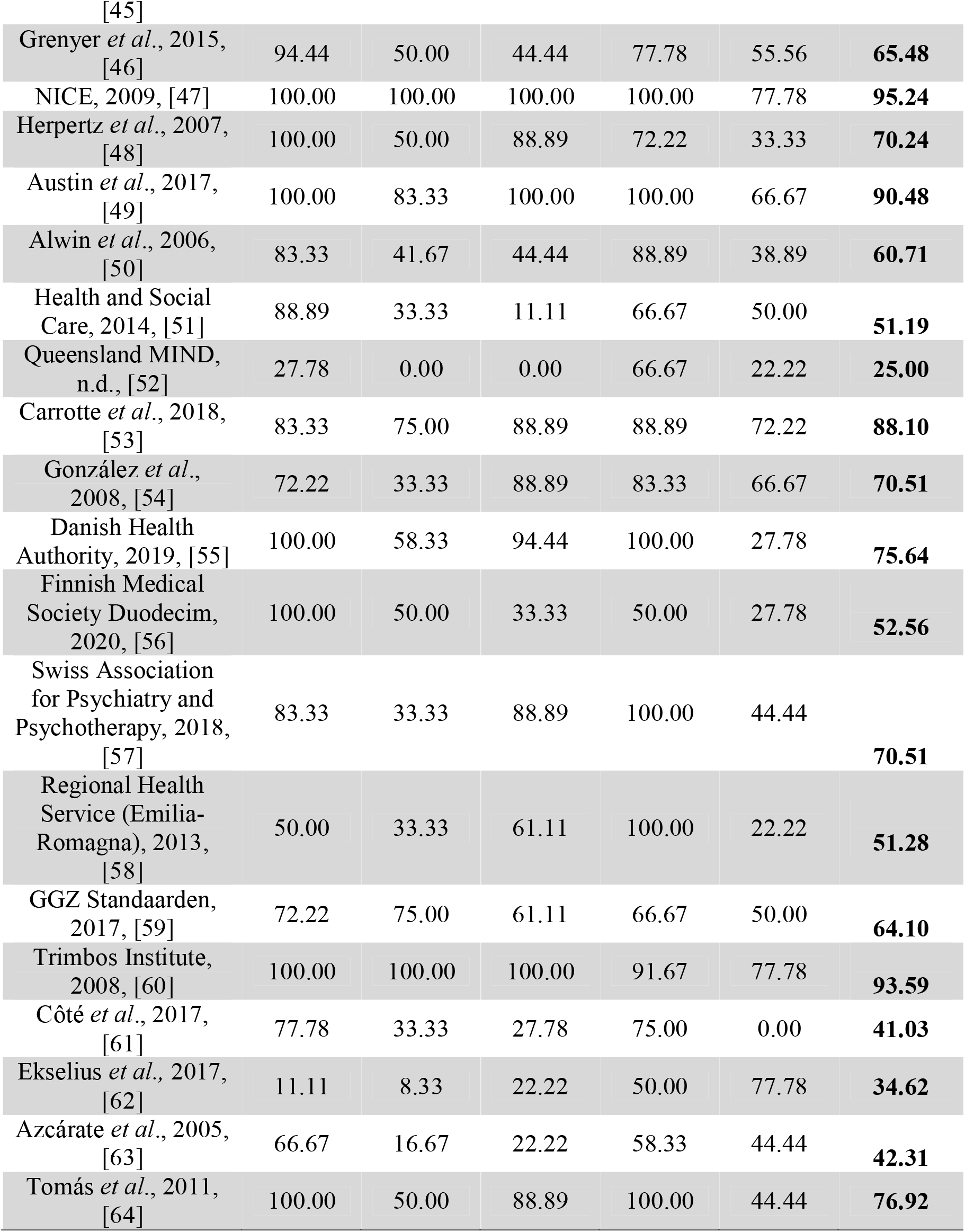
Individual domain and total scores of quality appraisal (AGREE-II).

Overall, Guidelines in this review performed the best in the following order (n= number of guidelines scoring as high quality in this domain): domain 1: Scope and purpose (n

= 22), domain 4: Clarity of presentation (n = 20), domain 3: Rigour of development (n = 11), domain 2: Stakeholder involvement (n = 9), and domain 5: Applicability (n = 8). Most of the guidelines had a clear purpose and rationale, with recommendations easily identified. Evidence considered that led to the recommendations were also well presented. However, very few guidelines had a clear methodology section that reported how evidence was gathered and subsequently analysed. This was reflected in the low number of guidelines assessed to be of high quality in domains 2, 3, and 5.

## Main Findings

We grouped findings from the guidelines into four organising domains: recipients of services, service delivery, staff, and treatment. Within each domain, more inductive themes are described. See Table 3. for a description of the themes and subthemes. A more detailed summary of themes and examples is provided in the Supporting Information S5.

**Table 3.**
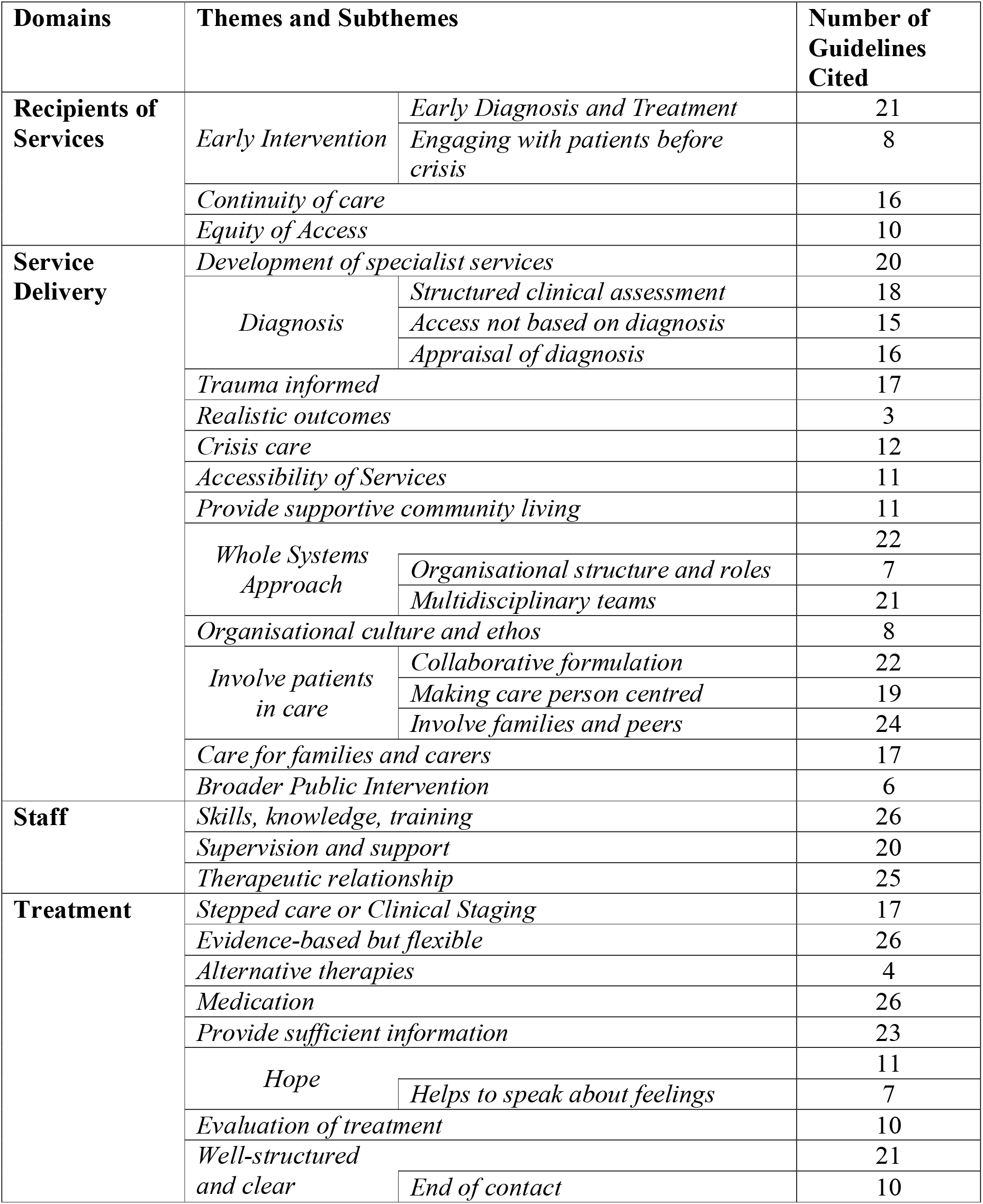
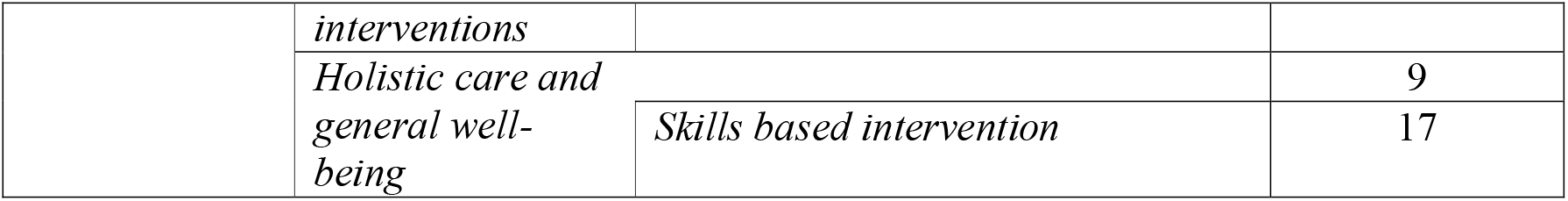
Detailed description of themes and subthemes.

### Recipients of services

The first domain looks at the provision of support, making recommendations about who should be receiving services and when such services should be provided. Thirteen guidelines stated that there should be *continuity of care*, where care is uninterrupted with planned transitions, support received from different services remains available to patients, and there is continuity between professionals. Guidelines suggested that services should be adequately equipped to serve different age groups, ensuring a smooth transition between services. This recommendation is not only limited to transitions related to age but also between other services such as forensics and general mental health services:

> *“There is also a need to provide a seamless transition between services that meet the needs of the client at each stage in their lives, moving through services for children to adolescent services to adult services to services for older adults*.*”*
>
> BPS: Understand Personality Disorder (UK)

Ten guidelines further stated that there should be *equity of access* where services provided are not influenced by one’s race, gender, faith, disability, or sexual orientation:

> *“Ensure that [individuals with CEN] belong[ing] to the ethnic minorities have equal conditions and opportunities for access to services culturally appropriate on the basis of clinical needs, through cultural mediation*.*”*
>
> Mental Health Department (Italy)

Twenty-one guidelines agreed that *early intervention* is of paramount importance, allowing treatment to be provided quickly upon diagnosis:

> “*Screening is intended to screen people suspected of having a personality disorder and early identification of those who qualify for more extensive diagnostics. The goal of this is to recognize the problem as soon as possible [and be able to offer] treatment at an early stage…”*
>
> GGZ: Standaarden (the Netherlands)

### Service delivery

All guidelines included in this review provided recommendations on how certain aspects of services should be delivered, from the quality of services to the service structure of hospitals and clinics. Twenty of the guidelines advocated the *development of specialist services* for the management of complex cases of CEN and provision of specialist training for other mental health services:

> *“Specialist teams should develop and provide training programmes that cover the diagnosis and management of borderline personality disorder and the implementation of this guideline for general mental health, social care, forensic and primary care providers and other professionals…”*
>
> NICE: BPD Treatment and Management (UK)

Most of the guidelines (n=16) made recommendations on the procedure of *diagnosing* CEN and the implications of a diagnosis. Diagnosis was seen as a gateway to care by some guidelines. However, eleven guidelines also cautioned the usage of stigmatising labels, warning that a diagnosis could bring more distress, negatively impacting their lives:

> *“The label ‘borderline personality disorder’ should be used with caution as it often has negative connotations (especially for health professionals) and may be associated with substantial stigma*.*”*
>
> COPE: Perinatal Mental Health Guide (Australia)

Eighteen guidelines agreed to adopt a *structured clinical assessment* to facilitate diagnosis. However, three international guidelines caution clinicians on the validity of translated diagnostic tools:

> *“In the international literature there are numerous psychometric tools for evaluation of [CEN]. However, only a part of these is currently translated and validated in Italian*.*”*
>
> Mental Health Department (Italy)

Seventeen guidelines made it clear that services should be *trauma informed* when making a diagnosis and formulating a treatment plan, ensuring that sensitive issues brought up were addressed:

> *“Health professionals need to be aware that many people with BPD have experienced significant trauma, either in the past or in their daily lives*.*”*
>
> National Health and Medical Research Council (Australia)

Eleven guidelines stated that *accessibility of services* should be a priority, with ready access to care not only within but also outside working hours. SANE Australia was the only guidelines that went a step further and advocated *digitalizing of services*, allowing more to receive treatment. However, in other settings such as the UK greater caution was suggested, with a need to evaluate whether digital platforms are effective in reaching more people:

> *“Digital platforms may offer opportunities to increase access in rural and remote settings… modifying treatments to suit digital platforms*.*”*
>
> SANE (Australia)

Twenty-one guidelines noted that the provision of services is a collective and multidisciplinary effort, hence suggested a *whole systems approach* where multiple services work together to provide an individual with effective care over time. Seven guidelines further suggested that within health care systems, clear roles should be specified to prevent conflicting responsibilities:

> *“Provision of services for people with ASPD often involves significant interagency working. Pathways between services should be clear, and communication between organisations should be effective*.*”*
>
> Meeting the challenge, Making a difference: Practitioner Guide (UK)

Most guidelines (n=19) recommended to *involve patients in their own care*, including *collaborative formulation* to ensure that all experiences of the patients are being addressed, allowing patients to develop autonomy over their own treatment:

> *“Services will promote personal decision making and help the individual build their capacity to manage their own mental health and wellbeing*.*”*
>
> HSC (Northern Ireland, UK)

Twenty-one guidelines also recommended that *families and carers* of patients should be involved throughout the recovery process, providing more information when diagnosing and being part of the treatment plan. Moreover, the emotional stress families and carers might experience should not be neglected:

> *“Provide family members and carers with information about the illness if appropriate, as well as reassure and validate their experiences with the person. Encourage family members and carers to look after themselves and seek support if required*.*”*
>
> MIND (Australia)

Lastly, six guidelines discussed having *interventions for the wider public*, educating the public on CEN to reduce existing stigma:

> *“Actively engage in mechanisms to bring about attitude change”*
>
> MIND Consensus statement (UK)

### Staff

Almost all the guidelines made recommendations on the skills and training that staff need, ensuring that they possess the competencies to provide the best possible care. Themes revolved around the provision of *training*, ensuring that staff have the right *skills* and *knowledge*:

> *“Diagnosis should be handled by qualified healthcare professionals who are trained in the use of recognized, valid and reliable diagnostic methods, and has familiarized himself with the manual of the diagnostic instrument, and has the opportunity to get supervision*.*”*
>
> Danish Health Authority (Denmark)

Staff were also expected by twenty-five guidelines to be skilled in forming a *therapeutic relationship* with patients, one that is filled with care and trust, allowing patients to have faith in the staff themselves as well as the recovery process. This was identified as crucial by the majority of the guidelines, and was seen as resulting in greater service engagement and treatment adherence:

> *“In order to engage these people in services it will be necessary to foster an attitude of respect for their suffering and an approach that recognises their dignity as fellow human beings*.*”*
>
> BPS: Understand Personality Disorder (UK)

Lastly, twenty guidelines recommended providing sufficient *support and supervision* to staff. Supervision and support in this context has two purposes: it is needed firstly to ensure staff can make the right diagnoses and deliver high quality treatment, and secondly to support the mental and physical well-being of the staff themselves:

> *“practitioners also need access to regular supervision. Without this there is likely to be a high degree of staff burn out, absenteeism, sickness and disillusion, and services may fail*.*”*
>
> PD: No longer a diagnosis of exclusion (UK)

### Treatment

The final domain revolves around the provision of treatment, with guidelines suggesting therapies of various types and making recommendations about how they should be delivered and structured. Seventeen of the guidelines recommend that services adopt a *stepped care model*, providing a framework for health care professionals to organise services and to identify the most effective interventions for the patient. Such a framework is intended to ensure that the limited resources are spent appropriately and that patients receive care matched to their needs:

> *“stepped care approach is used to match their needs with the right level of support; the individual only ‘steps up’ to intensive / specialist services as their needs require*.*”*
>
> HSC: Regional Care Pathway for PD (Ireland)

Almost all guidelines (n=26) agreed that treatments administered must be backed by evidence, and patients should have the *flexibility* of selecting the treatment that they would want to receive. Moreover, interventions should be clearly explained to patients, providing them with *sufficient information*. However, guideline from the Netherlands cautioned users to not completely dismiss therapies that have yet to be backed by evidence, given that research in this area is in its infancy:

> *“Others forms of treatment … model are promising and serve to be further investigated for their effectiveness. They certainly do not need to be discouraged at this point or excluded*.*”*
>
> Trimbos Institute (the Netherlands)

Interestingly, art-based therapies such as art, drama, and music were not recommended by NICE as they lack sufficient evidence, however, guidelines from Spain and the Netherlands recommend them as an *additional therapy*, on top of existing treatments:

> *“[alternative] therapies become seen as an addition to psychotherapeutic treatment, among other things to gain access to emotions in patients who are (emotionally) difficult to reach*.*”*
>
> GGZ: Standaarden (the Netherlands)

Adopting a *well-structured intervention* was recommended by twenty-one guidelines, as well as ensuring that treatments are being administered as intended, and that *routine evaluation* allows swift identification of ineffective treatment. Only ten guidelines recommended clinicians to discuss the *end of contact* with therapy with patients, ensuring that patients do not experience a sudden loss of support:

> *“use of competence frameworks based on relevant treatment manuals, routine use of sessional outcome measures, routine direct monitoring and evaluation of staff adherence”*
>
> NICE: Anti-social PD Treatment and Management (UK)

Apart from focusing on the defining features of CEN, nine guidelines advocated *holistic care* for the patients, ensuring that their *general well-being* is taken care of. This includes having sufficient rest and nutrition:

> *“work for health promotion, where one tries to influence the individuals’ lifestyle and behaviour to promote health…”*
>
> Swedish Psychiatric Association (Sweden)

Seventeen guidelines also recommended signposting individuals to services that could support them in developing important functional and occupational skills, as well as addressing social needs such as dealing with loneliness and housing:

> *“Services will value the individual as a person and help them develop a positive and solution focused approach to the management of their needs. Services will work to enable the individual to maximise their personal strengths, resources and talents*.*”*
>
> HSC: Regional Care Pathway for PD (Ireland)

## Discussion

The aim of this systematic review was to identify recommendations made by different organisations from across the world on community treatment for people with CEN. We identified four main domains, with a total of 27 themes after synthesising 29 guidelines from 12 different countries and organisations. Themes identified related to the provision of services, how services differed according to life stage and clinical needs of individuals with CEN, how and when services should be delivered, staffing and training required for these services, and the delivery of treatments.

A common statement made across the majority of the guidelines, was that research to support the formulation of clinical recommendations is lacking. Despite this lack of evidence, mental health professionals still have the responsibility to treat patients as effectively as they can. To support them in doing this, a body of guidance has been developed that is to a large extent based on expert consensus about best practice. Trial evidence was available to support some recommendations, but most guidelines were rated as low quality in relation to use of evidence to support recommendations.

The overall quality of included guidelines varied considerably. Unsurprisingly, authors who adopted a quality appraisal tool as a methodological strategy for the development of the guideline obtained higher scores in all five domains (for example guidelines from NICE where the AGREE framework was implemented throughout the guidance). AGREE is intended as an international tool for any authors to utilise and not just available to UK based guidelines. We also observed that guidelines with higher scores tended to also be those with clinicians as their target audience. As observed from Table 1, most guidelines were labelled as ‘yes’ for being evidence-based and having involved service users. However, this was not reflected in domains 2 (stakeholder involvement) and 3 (rigour of development) of the quality appraisal, with most of the guidelines performing poorly. This mismatch is due to guidelines stating that recommendations were based on evidence and that service users were involved in their development, but failing to elaborate on how this was achieved.

Unlike the previous review by ESSPD [23], the current review included guidelines beyond the European region, including North America and Oceania. Our findings are in line with the three main areas reported (diagnosis, psychotherapy, and pharmacotherapy) in the previous review by ESSPD [23]. Both this review and the review by ESSPD found that most guidelines recommended the following: to adopt a structured clinical assessment when making a diagnosis, abstain from pharmacological therapy due to insufficient evidence to support its efficacy, and employ psychological therapies that are backed by clinical evidence. This is not surprising as seven out of nine of the guidelines included in the ESSPD review were also included in this systematic review. The current review additionally identified recommendations made on service structure, provision of staff and treatments targeted at general well-being.

The themes of this systematic review are in line with those identified in a recent co-produced qualitative interview study by Trevillion *et al*. [15] which included in depth interviews with 30 individuals with CEN and a qualitative thematic meta-synthesis by Sheridan Rains *et al*. [27] which looked at the needs of 1531 service users with CEN. Clinicians were perceived by service users to have expectations of swift recovery; this mismatch between expectations and reality could deter service users from engaging with staff [27]. Both Trevillion *et al*. and Sheridan Rains *et al*., [15, 27] also concluded that most of the participants reported negative experiences with services. These conclusions emphasise the importance of having a good fit between what service users identify as priorities and the recommendations made in guidelines. Trevillion *et al*. [27] only recruited participants from the UK so findings can only be extrapolated to UK services, and further research into service users’ experiences in other countries is warranted. Nevertheless, the guidelines produced in the UK did include recommendations that addressed the majority of the needs stated by services users in the Trevillion et al. [15] and Sheridan Rains et al., [27] studies suggesting that the issue does not lie within the recommendations of guidelines but the implementation of these recommendations in services.

From a different perspective, Foye *et al*. [16] conducted a qualitative study with 50 clinicians and Troup *et al*. [28] conducted a systematic review and qualitative thematic meta-synthesis of published research with a total of 550 clinicians, both exploring clinicians’ perspectives of best practice community care for individuals with CEN. Similarly, most ofthe themes identified were in line with the themes of this systematic review as well as service users’ needs, reinforcing the notion that current guidelines are a good fit with the priorities identified both by services users and clinicians. The need for holistic care that goes beyond treating symptoms of CEN was, however, lacking in most of the guidelines, with only nine guidelines in this systematic review giving recommendations to provide holistic care, looking beyond the medical needs of individuals with CEN.

Dissemination and implementation plans of the included guidelines were limited; of the twenty-nine guidelines in this review only eight included such a plan. This might explain the gap between why existing guidelines contained similar recommendations to meet service users’ needs as identified in other recent qualitative research, yet service users are still not experiencing the quality of care set out by these guidelines. Moreover, guidelines were not mentioned by clinicians in the meta-synthesis by Troup et al. [28] to be of either help or no help in treating and managing individuals with CEN, suggesting that guidelines might not actually be routinely consulted or are not known by these clinicians.

## Strengths and Limitations

To our knowledge, this systematic review is one of the first to explore the consensus between international guidelines on recommendations for community-based treatment for people with CEN. However, using google translate to extract information from guidelines written in languages other than English means that there is a risk of missing potentially relevant points in some sections of guidelines which may have been poorly translated.

Although, the number of guidelines included in this review is thrice as many as the previous ESSPD systematic review [23], including guidelines from North America, Western Europe, and Oceania, the cultural composition of the included countries is still western centric. Guidelines that are from the African or Asian region might contain relevant materials for this systematic review. Including guidelines that are culturally diverse would allow themes to be more culturally sensitive and provide recommendations that might work better for people from different cultures.

### Implications for Policy and Practice

The findings of this systematic review highlight the common recommendations in existing international guidelines with regards to the provision of community services for CEN. These guidelines seem generally congruent with the priorities that service users and clinicians identify, hence it is helpful for clinicians treating people with CEN to be aware of these guidelines and consult them regularly. However, as observed from this systematic review, many recommendations are not backed by research evidence, further highlighting the need for good quality research into the community treatment of CEN. Lastly, different practices recommended in the guidelines of different countries highlights wider possibilities for the treatment of CEN, which could be considered by clinicians and researchers.

### Future Research

Future research could helpfully explore potential barriers that are preventing professionals from consulting guidelines in practice. Governing bodies should investigate fidelity to guidelines and the degree to which current guidance is being adhered to in actual practice. Fundamentally, more high-quality research into the treatment of CEN is required in order to shape future guidelines. A future review can be conducted to include these changes and assess if the needs of service users are being met according to the updated guidelines.

### Conclusion

This systematic review synthesised recommendations made from various international guidelines with regards to the provision of services for individuals with CEN in a community setting. It is apparent that existing guidelines have the potential to support clinicians in meeting many service users’ needs. There was consensus on a set of priorities for good practice that seem to have some congruence with sources on services users’ and clinicians’ priorities, in at least some areas. However, it is not clear to what extent guidelines are being adhered to in practice. Half of the current guidelines were of lower methodological quality, with many recommendations not backed by evidence, highlighting the urgent need for more high-quality research into the treatment of CEN and further guideline development and dissemination.

## Data Availability

All data is already available in the public domain.

## Supporting Information

**S1 Search Strategy of bibliographic databases**. This includes the complete search strategy used in each bibliographic database and an example strategy of the original search.

**S2 Search strategy of guideline databases**. This includes the full list of guideline databases and organisations included in this search.

**S3 Search terms in different languages**. This includes search terms used in the additional eight languages we searched in addition to English.

**S4 Amendments made to the Appraisal of Guidelines for Research and Evaluation II framework (AGREE-II)**. Amendments were made based on the appropriateness of the question to the studies aims.

**S5 Detailed summary of themes**. This includes a more detailed summary of themes extracted from the guidelines with quotes.

