## Supporting Information 5 for "Evaluation of international guidance for the community treatment of complex emotional needs: A systematic review"

**Supplementary Material 5. Long Summary of Findings**

| **Theme** | **Description** | **Citing Guidelines** |
| --- | --- | --- |
| **Recipients of Services** | Recommendations of who should have access to services, and when. |  |
| **Early Intervention** | Several guidelines highlighted the importance of intervening early in the treatment of personality disorder. These recommendations tended to centre around: |  |
| *Continuity of care* | Guidelines mentioned the transition of individuals diagnosed with complex emotional needs from adolescence to adulthood should still be experiencing the same level of care when transiting towards a new service or organisation.  *There is also a need to provide a seamless transition between services that meet the needs of the client at each stage in their lives, moving through services for children to adolescent services to adult services to services for older adults.*  BPS: Understand Personality Disorder (UK)  This transition is not only limited to age groups but also between other services such as between forensic and general mental health  *Clear protocols would need to be drawn up to set out the mechanics for the transfer of the care of patients between general mental health and forensic services.*  PD: No longer a diagnosis of exclusion (UK) | 13 |
| *Early Diagnosis and Treatment* | Several guidelines also highlighted the importance of intervening early, and ensuring that services are adequately prepared to diagnose and treat  *The critical importance of childhood and adolescence in setting the course for a healthy adult life make it essential that early signs are recognised and effectively addressed.*  MIND Consensus statement (UK)  *health professionals should make the diagnosis of BPD in adolescents and young people who meet diagnostic criteria, so that early intervention can begin without unnecessary delay*  National Health and Medical Research Council (Australia)  *Screening is intended to screen people suspected of having a personality disorder and early identiﬁcation of those who qualify for more extensive diagnostics. The goal of this is to recognize the problem as soon as possible and so treatment at an early stage to be able to offer.*  GGZ: Standaarden (Netherlands) | 21 |
| *Engage with patients before crisis* | Care should be provided before patient’s reach crisis point  *It is recommended to identify the factors that precipitate or exacerbate the [symptoms] and promote early interventions to prevent relapse.*  Departament de Salut (Spain) | 8 |
| **Support throughout the lifespan** | A few guidelines noted that services provided to patients should be equipped to serve older adults, who might face further complications as they age  *Particular attention needs to be paid to older people to ensure that their needs are not forgotten and are properly understood and further research needs to be undertaken to understand this group of people better.*  MIND Consensus statement (UK)  *In the elderly, the diagnosis of BPD can be complicated by the table potentially altered clinic. When the classic acute symptomatology of BPD is absent, However, we should not give up making a diagnosis. Lack of recommendations empirically validated speciﬁc treatments necessitates the support of treatments speciﬁc to the disorder by general recommendations for geriatric psychotherapy.*  SAPP: Treatment recommendations for BPD(Switzerland) | 5 |
| **Service Delivery** | Recommendations of how services should be delivered forms the bulk of most guidelines. A total of 12 different service delivery components were identified from all the guidelines: |  |
| **Development of Specialist services** | Most of the guidelines recommended the creation of specialist services for complex emotional needs, and tasking this service with educating the wider public health system as alongside treating complex cases  *Some clinicians have suggested that services for those with personality disorder require highly specialised skills and need to be developed as separate – or tertiary – services; there are problems in engaging those with personality disorder in treatment and specialised dedicated services may be better at this.*  BPS: Understand Personality Disorder (UK)  *The systematic development of the treatment of unstable personality requires the establishment of special working groups with sufﬁcient skills and effective professional development. to ensure in-service training.*  FMSD: Unstable Personality (Finland)  *Specialist teams should develop and provide training programmes that cover the diagnosis and management of borderline personality disorder and the implementation of this guideline for general mental health, social care, forensic and primary care providers and other professionals who have contact with people with borderline personality disorder.*  NICE: BPD Treatment and Management (UK) | 20 |
| **Crisis Care** | Guideline recommended formulating a crisis care plan early on, allowing patients and their carers to react timely and safely  *It is very important for family, partners and immediate others that care providers realize what a huge impact a crisis situation has on both the patient and his immediate environment. For that reason, it is very important that crisis situations are properly assessed from the zero line and that the necessary guidance is provided receives.*  TRIMBOS Institute (Netherlands)  *If someone is sensitive to a crisis, a crisis plan can be drawn up with the patient and, if possible, with the next of kin be worked out so that all involved know what to do in the event of a crisis. When the one ready is, the plan can be elaborated into a signalling plan, so that a crisis can be prevented or becomes less violent.*  GGZ: Standaarden (Netherlands)  Patients usually present themselves in times of crisis, hence hospitals should be prepared and equipped to manage such situations  *The hospital crisis intervention offers the possibility to patients with a deﬁcit in terms of self-control and a high risk of endangerment of control and support, for example when suicidal attacks.*  *~*  *At most, a hospital crisis intervention should last from a few days to 2 weeks and not should not take place in a closed acute psychiatric ward but in an open crisis ward. To the extent that structural characteristics allow (for example monitoring narrow), an open therapeutic framework is also recommended in acute suicidality. Ideally, admission should take place on a voluntary basis. Coercive measures such as isolations or even ﬁxings should be absolutely avoided.*  SAPP: Treatment recommendations for BPD(Switzerland) | 12 |
| **Equity of Access** | Some guidelines also mentioned that access to services should not be limited to one’s race, recommending solutions to treat patients of different cultural backgrounds  *People from Black Minority Ethnic groups should have access to culturally appropriate Personality Disorder Services based on their clinical need.*  HSC (Ireland)  *Ensure that people with Borderline Personality Disorder (PGD) belong to the ethnic minorities have equal conditions and opportunities for access to services culturally appropriate on the basis of clinical needs, through cultural mediation.*  Mental Health Department (Italy)  Apart from providing equitable services, some guidelines recommended a more aggressive stance in identifying patients who are harder to reach  *Services should adopt systems to identify patients with whom they are struggling to engage and promote engagement*  RCPsych Position Statement (UK)  *Comprehensive provision also means services that engage and support hard-to-reach population groups, and those with particular needs who may be overlooked. It is important that the development of PD services does not reinforce health and social inequalities.*  Recognising Complexity (UK) | 9 |
| **Diagnosis** | A majority of the guidelines gave recommendations of diagnosing complex emotional needs, this included implication of a diagnosis. Five subthemes were identified: |  |
| *Access not based on diagnosis* | A diagnosis should not be the main factor limiting or enabling a patient access to services  *There is a general agreement that diagnosis alone is insufficient and good care should be guided by a co-constructed biopsychosocial formulation which gives patients an experience of being understood. A diagnosis should only be made after appropriately skilled and thorough assessment, although this should not cause a delay in receiving suitable interventions and care.*  RCPsych Position Statement (UK)  *For example, telling a person their behaviour is not severe enough to be admitted to the inpatient unit may actually increase self-harming behaviours to gain hospital admission.*  Illawarra Health and Medical Research Institute (Australia)  *it is vital to recognise that need for treatment may not be dependent on diagnosis.*  MIND Consensus statement (UK) | 16 |
| *Benefits of diagnosis* | Some guidelines did however mention that diagnosis could facilitate care for some patients  *When the diagnosis of BPD has been made, GPs can provide or support effective BPD treatment by ensuring that other health professionals within the service are aware of the general principles of caring for a person with BPD*  National Health and Medical Research Council (Australia)  *Discussion of the formulation can help the individual (and family/ carers where appropriate) understand their needs and help identify areas of their lives which may need to change.*  HSC (Ireland) | 11 |
| *Not using stigmatising labels* | However, for some patients, a diagnosis would bring more distress, negatively impacting their lives. Some guidelines address this by providing solutions to address this stigma  *The term ‘borderline’ is not meaningful to people with BPD and their families and friends and, for some people, it may have associations with blame and stigma. Therefore, the clinician should explain the condition in a sensitive, non-judgemental way that conveys that it is not the person’s own fault, but a condition of the brain and mind that is associated with both genetic and environmental risk factors.*  National Health and Medical Research Council (Australia)  *The label ‘borderline personality disorder’ should be used with caution as it often has negative connotations (especially for health professionals) and may be associated with substantial stigma.*  COPE: Perinatal Mental Health Guide (Australia) | 11 |
| *Structured clinical assessment* | Among the different forms of assessing complex emotional needs, some guidelines recommended structured clinical assessments, allowing patients to receive the necessary support  *Structured assessments are essential to services treating individuals with the problems of personality disorder.*  BPS: Understand Personality Disorder (UK)  *Staff involved in the assessment of antisocial personality disorder in secondary and specialist services should use structured assessment methods whenever possible to increase the validity of the assessment.*  NICE: Anti-Social PD Treatment and management (UK) | 18 |
| *Adaption of diagnostic tools* | Some guidelines from countries where English is not their native language mentioned the validity of instruments after being translated and adapted to fit the local context  *Many of the instruments used in the assessment of personality disorders they have been translated, adapted and validated in our environment.*  Departament de Salut (Spain)  *In the international literature there are numerous psychometric tools for evaluation of DGPs. However, only a part of these is currently translated and validated in Italian.*  Mental Health Department (Italy)  Apart from language differences, a guideline also pointed out cultural differences  *Use appropriately translated versions of the EPDS with culturally relevant cut-off scores. Consider language and cultural appropriateness of the tool.*  COPE: Perinatal Mental Health Guide (Australia) | 3 |
| **Realistic Outcomes** | A few guidelines noted that clinicians should set realistic goals of treatment, incentivising treatment engagement through smaller attainable goals  *An important point is that people with personality disorders should be careful not to ask too much. Psychological problems and treatment can demand a lot from someone. When asking goals, such as exercising more or getting ﬁnances in order, it is important that these realistically as possible. It can help the person plan to achieve the goal divided into small steps. It is good if one is kind to oneself, oneself de gives room to make mistakes and does not see the failure or only partial achievement of goals as a evidence of failure and failure.*  GGZ: Standaarden (Netherlands) | 3 |
| **Trauma Informed** | Being aware of patients past traumatic experiences was noted by a few guidelines to better assist in diagnosing as well as creating a treatment plan that would be beneficial for the patient  *Health professionals need to be aware that many people with BPD have experienced significant trauma, either in the past or in their daily lives. A high proportion of people with BPD report physical or sexual abuse or neglect during childhood.*  National Health and Medical Research Council (Australia)  *always consider, when activating a treatment, that many people may have experienced rejection, abuse, trauma and may have experienced stigma often associated with self-harm and the diagnosis of Personality Disorder.*  Mental Health Department (Italy) | 17 |
| **Accessibility of services** | Multiple guidelines mentioned providing adequate services to support the needs of patients, this includes access to services at non-working hours  *In acute situations, the patient must be able to count on 24/7 availability and accessibility from practitioners. The following general rules apply: the Treek standards apply for waiting times; the care location / practice is easily accessible by car and public transport*  GGZ: Standaarden (Netherlands)  and also, the accessibility of the hospital or clinic  *Increasing availability of evidence-based treatment services in rural, regional and remote areas. This could be achieved through a number of strategies, including: Funding dedicated specialist services in rural settings, such as major rural cities. Where it is not feasible to implement a specialist service, consider implementing evidence-based brief interventions. Increasing funding current training services to travel to these areas and conduct outreach training programs focusing on personality disorder treatment principles and evidence-based treatment methods such as DBT and schema therapy. Reviewing the efficacy of current incentives to attract clinicians to rural and remote areas.*  SANE (Australia) | 11 |
| *Digitalising services* | One guideline recommended shifting components of the service online, allowing patients to have better access to services  *Developing and funding evidence-based digital psychotherapy options. Digital platforms may offer opportunities to increase access in rural and remote settings. Consider increasing digital and phone-based options and modifying treatments to suit digital platforms. While telepsychiatry has proved promising in the treatment of some illnesses, comprehensive specialist programs are not always feasibly conducted via Skype or other online platforms. For example, it is quite difficult to provide group skills training online in real time. Hence, work in this field would need to involve a review of existing evidence and the feasibility of conducting such efforts in Australian settings.*  SANE (Australia) | 1 |
| *Provide supportive community living* | There was also a call for services to extend beyond the clinical setting, providing support in the community, hence providing patients with better a transition  *A coordinated ongoing community treatment model, which supports continuity of care and is understood within a relational model, is essential to the effective treatment of personality disorders. This is to prevent experiences of treatment as limited, fragmented and episodic.*  Illawarra Health and Medical Research Institute (Australia)  *The lack of adequate community-based provision has a triple effect. The endless cycle of rejection can intensify the distress and therefore the difficult behaviour of some individuals. It also means that those who are receiving intensive support and therapy, possibly in in-patient settings, can experience numerous barriers in returning to the community and in coping*  PD capabilities framework (UK) | 11 |
| **Whole Systems Approach** | Finally, multiple guidelines suggested that the provision of services does not fall solely on a single department within the system, but multiple teams and department working together  *It is arguable that outcomes for people receiving a diagnosis of personality disorder substantially depend on system behaviour across multiple service domains such as children’s services, education, social care, criminal justice, public health, public policy and primary care.*  MIND Consensus Statement (UK)  *It is therefore vital that strong links exist across these organisations to ensure effective care is provided. In addition to health and the criminal justice system, housing, adult education and the voluntary sector services will be required.*  NICE: Anti-Social PD Treatment and Management (UK) | 8 |
| *Case management* | A couple of guidelines included recommendations on how cases should be managed between services  *Regular communication between those who providing performance and treatment to patients is needed to ensure consistency and effectiveness.*  Mental Health Department (Italy)  *A coordinating practitioner is responsible for the treatment process and for adequate communication with other involved practitioners and service providers.*  GGZ: Standaarden (Netherlands) | 19 |
| *Organisational structure and roles* | On a similar note, some guidelines recommended having specific roles of services and individuals clearly defined, preventing conflicts within the health care system  *In addition to clarity about the task and values of the service, it helps if there is equal clarity about people’s roles and responsibilities.*  Meeting the challenge, Making a difference: Practitioner Guide (UK)  furthermore, having a competent leader to lead the team was also noted in a guideline  *Team leaders can play an important role in modelling adaptive and constructive responses to people with personality disorder. Support from management is also vital in acknowledging that clinical supervision and consultation is a legitimate part of the clinician’s role and does not signify weakness or ineffectiveness.*  Illawarra Health and Medical Research Institute (Australia) | 7 |
| *Multidisciplinary teams* | Guidelines also recommended having teams from different disciplines to provide better support for patients  *Especially in the treatment of patients with complex or severe personality disorders, and where it is otherwise estimated that there is a need for multidisciplinary competencies,*  Danish Health Authority (Denmark)  *People with personality disorder need a multidisciplinary and multiagency service.*  BPS: Understand Personality Disorder (UK)  *Provision of services for people with ASPD often involves significant interagency working. Pathways between services should be clear, and communication between organisations should be effective.*  Meeting the challenge, Making a difference: Practitioner Guide (UK) | 21 |
| **Organisational Culture and Ethos** | Recommendations to slightly alter the cultural environment of the hospital, making it more conducive for recovery was noted by a few guidelines, it is hope that such changes would allow patients to be more engaged in treatment  *To create this environment of mutual respect, tolerance, understanding, and availability for change, common rules of the game are needed, respected by all.*  Ministry of Health and Consumption (Spain)  *Having access to their creativity can then be crucial edge. And creativity grows best in an environment characterized by joy and a measure of playfulness.*  Swedish Psychiatric Association (Sweden) | 8 |
| **Involve patients in care** | A number of recommendations were made around involving patients in their own care, these centred around five sub-themes: |  |
| *Collaborative formulation* | There was a clear consensus that formulation of the problems experienced by the patient should be developed in collaboration with the patient, so that they can ensure all aspects of their experience are being addressed.  *Be treated as a partner in the delivery of the service and supported to work collaboratively towards agreed goals.*  HSC (Ireland)  *Particularly important is a clear psychological formulation underpinning the service plan which is produced in collaboration with the individual receiving care*  MIND Consensus statement (UK)  *It is recommended to involve and commit the patient in their formalized treatment in the establishment of a contract that speciﬁes the therapeutic framework and the responsibilities of professionals, patient and family, if applicable.*  Departament de Salut (Spain) | 16 (15) |
| *Developing autonomy* | Guidelines also recommended methods of ensuring patients autonomy throughout the treatment process, encouraging engagement  *Services will promote personal decision making and help the individual build their capacity to manage their own mental health and wellbeing.*  HSC (Ireland)  *While people with Borderline Personality Disorder (PGD) can do enormously struggles in making informed and aware choices, especially in crisis phases, the efforts made by professionals to induce the solutions they consider best can limit development self-efﬁcacy. For this reason it is necessary to encourage patients and their families a think about the different options of choice and reﬂect on the consequences of each of them.*  Mental Health Department (Italy) | 19 |
| *Making care person-centred* | Along a similar line, it was also stressed that care should be person centred, as each patient brings their own unique presentation  *Patients seldom come to assistance with complaints that directly affect their personality. They usually want to be helped in connection with complaints such as sadness, anxiety, sleeping problems or relationship problems. In many cases, it is necessary to institute a complaint-oriented treatment, in accordance with the request for help of the patient,*  TRIMBOS Institute (Netherlands)  *Care coordination is a collaborative process of assessment, planning, facilitation and advocacy to meet an individual’s mental health needs through communication and available resources to promote the best possible outcome of the consumer*  Illawarra Health and Medical Research Institute (Australia)  *The focus was to see the whole person in their social and historical context, not just the disease*  Swedish Psychiatric Association (Sweden) | 20 |
| *Involve families* | A majority of the guidelines also noted methods of engaging with the patient’s family during the diagnosis and treatment phase  *Family members are usually grateful to be educated about the borderline diagnosis, the likely prognosis, reasonable expectations from treatment, and how they can contribute. Such interventions often improve communication, decrease alienation, and relieve family burdens.*  A BPD Brief (USA)  *In addition, it may be helpful where possible, and with the consent of the individual, to obtain information from family members and friends. This can enrich the assessment process and provide an alternative perspective that can corroborate or challenge the individual’s presentation of their problems.*  BPS: Understand Personality Disorder (UK) | 22 |
| *Peer support* | Similarly, a few guidelines asserted that patients interacting with others who are or were in a similar position would be beneficial for recovery  *Patients meet other patients, which can provide an opportunity to experience other ways of being in relation to other people*  Danish Health Authority (Denmark)  *Peer support/social activities. Spending time with others who share similar experiences can be immensely powerful in generating hope, learning new coping strategies and making connections with others.*  Meeting the challenge, Making a difference: Practitioner Guide (UK) | 8 |
| **Care for families, and carers** | Several guidelines looked beyond providing care only for the patients, care for the patient’s families and carers were also recommended  *Those providing regular and substantial care will be offered an assessment of their own needs which is reviewed regularly*  HSC: Regional Care Pathway for PD (Ireland)  *Provide family members and carers with information about the illness if appropriate, as well as reassure and validate their experiences with the person. Encourage family members and carers to look after themselves and seek support if required.*  MIND (Australia)  Patients with children were also of concern to some organisations, with a few guidelines recommending clinicians to address the needs of such children  *The needs of dependents, especially their possible dependent children*  Departament de Salut (Spain)  *inform children of parents with a personality disorder about the problem and check in the extent to which the child experiences problems (parentiﬁcation, detachment, problems at school, etc.) and whether support is required for this*  GGZ: Standaarden (Netherlands) | 17 |
| **Broader public intervention** | A few guidelines mentioned interventions that targets a wider audience, educating the general public on complex emotional needs and to ensure that this shift in perception goes beyond the hospital setting  *Actively engage in mechanisms to bring about attitude change i.e media campaigns*  MIND Consensus statement (UK)  *Design and conduct formative research into a multi-channel media campaign aiming to educate the Australian community and destigmatise personality disorder.*  ~  *There is a need to improve awareness of personality disorder within the general community, including teachers and others working in schools. A significant component of this campaign may also involve ‘myth-busting’. Any such initiatives will need to be carefully designed, and to involve individuals with personality disorder, and carers, families and other support persons, throughout development (Grenyer, 2017), as well as input from health professionals and researchers.*  SANE (Australia)  *implement public campaigns for “awareness of mental health*  Departament de Salut (Spain) | 6 |
| **Staff** | A majority of the recommendations pointed out how to best utilise staff, ensuring that they are sufficiently equipped to provide patients with the best possible care. Recommendations were based on three subthemes |  |
| **Skills, Knowledge, Training** | Most of the guidelines recommended that adequate training in diagnosis and treatment of complex emotional needs should be provided to all staff  *Diagnosing personality disorders is challenging. Diagnosis should be handled by qualiﬁed healthcare professionals who are trained in the use of recognized, valid and reliable diagnostic methods, and has familiarized himself with the manual of the diagnostic instrument, and has the opportunity to get supervision.*  Danish Health Authority (Denmark)  Furthermore, treatment should be administered based on the clinician’s proficiency  *From among the effective BPD treatments, therapists should offer the treatment approach that best matches their training, theoretical framework and preferences. The effectiveness of a psychotherapy may depend on the individual therapist, and not all therapists will achieve the same results with a particular therapy*  National Health and Medical Research Council (Australia) | 27 |
| **Supervision and support** | Apart from providing the best possible care towards patients, most guidelines also mentioned providing sufficient support for health care workers themselves. Supervision and support can be seen as providing guidance in diagnosing and treatment of personality disorder  *Service managers and team leaders are responsible for ensuring that caseloads for clinicians who treat people with BPD are appropriate and realistic according to the clinician’s experience, the needs of individuals according to phase of treatment, the requirements of the specific treatments provided, and the number of complex cases.*  National Health and Medical Research Council (Australia)  *This initial training needs to be followed by regular clinical supervision to reinforce the training and provide an environment where potential problems can be discussed,*  BPS: Understand Personality Disorder (UK)  Support is also seen as support provided towards healthcare workers ensuring that their physical and mental well-being are not neglected  *Some nurses report experiencing anxiety due to the unpredictable, stressful or apparently manipulative behaviour associated with some of the personality disorders.*  MIND (Australia)  *As well as having the appropriate personal characteristics to work effectively in this field, practitioners also need access to regular supervision. Without this there is likely to be a high degree of staff burn out, absenteeism, sickness and disillusion, and services may fail.*  PD: No longer a diagnosis of exclusion | 21 |
| **Therapeutic Relationship** | Clinicians were recommended to be skilled at forming a relationship filled with care and trust with patients, allowing patients to develop trust hence ensuring engagement with treatments  *Health professionals and other staff should encourage trust by showing a non-judgemental attitude and by being consistent and reliable (e.g. by keeping appointment times, making arrangements for contact outside the planned appointment schedule that are feasible to sustain long-term, planning for staff continuity over time, and explaining the team structure and team members’ roles to the person).*  National Health and Medical Research Council (Australia)  *In order to engage these people in services it will be necessary to foster an attitude of respect for their suffering and an approach that recognises their dignity as fellow human beings.*  BPS: Understand Personality Disorder (UK) | 25 |
| **Treatment** | Treatment recommendations were made by almost all the guidelines, treatment can be further broken down into 9 different aspects: |  |
| **Evidence-based but flexible** | Most guidelines agreed that treatment should be administered based on evidence, however flexibility should still be practiced, offering patients choices  *There is evidence that certain individual treatments do help (for example mentalization based therapy, schema therapy, and dialectical behaviour therapy) and for an emerging stepped care approach to treatment 35 but no one method appears to confer an advantage over another.*  MIND Consensus Statement (UK)  *DBT is effective in treatment of borderline personality disorder, with effects including a decrease in inappropriate anger, a reduction in self-harm and an improvement in general functioning (Stoffers et al 2012). While other treatments have been less evaluated, overall findings support a substantial role for psychotherapy in treating borderline personality disorder.*  COPE: Perinatal Mental Health Guide (Australia)  *In the multimodal treatment program, in addition to psychotherapy, one or more treatment approaches may be included, such as involves a signiﬁcant increase in the total treatment time (dose). Examples of such approaches are: Psychoeducation, mindfulness, body therapy or social skills training. It is a good clinical standard to continuously assess psychotherapeutic treatment courses and through this assessment clarify what type of treatment course the patient should be offered. Furthermore, it is important to choose the right level of treatment, neither too much nor too little.*  Danish Health Authority (Denmark) | 27 |
| **Additional Therapies** | A handful of guidelines discussed the possibility of incorporating interventions that are less explored, these included art, music, and drama therapy  *Professional therapies become seen as an addition to psychotherapeutic treatment, among other things to gain access to emotions in patients who are (emotionally) difﬁcult to reach. Also, some patients verbally less skilled or they cannot express their own emotions well with words, which means that in psychotherapy can be less focused on change. Professional therapy can be done here to offer a solution.*  GGZ: Standaarden (Netherlands)  *In the opinion of the working group (group) music therapy can be an important element in treatment programs for patients with personality disorders.*  TRIMBOS Institute (Netherlands)  However, some guidelines cited the lack of evidence to prove the efficacy of such therapy, hence not recommending it  *There is very little research on the effectiveness of arts therapies for people with borderline personality disorder and therefore no recommendations could be made.*  NICE: BPD Treatment and Management (UK) | 4 |
| **Medication** | Majority of guidelines discussed the possibility of including pharmacotherapy within the treatment plan, with most recommending it only to treat comorbidities and not personality disorder on its own  *Patients with BPD should be informed that there is no strong evidence base for the prescription of any drug. However, the off-label use of psychotropic agents may help individuals with BPD to improve affective symptoms and impulsivity.*  World Federation of Societies of Biological Psychiatry    *Pharmacological interventions for comorbid mental disorders, in particular depression and anxiety, should be in line with recommendations in the relevant NICE clinical guideline. When starting and reviewing medication for comorbid mental disorders, pay particular attention to issues of adherence and the risks of misuse or overdose.*  NICE: Anti-social PD Treatment and Management (UK) | 27 |
| **Provide sufficient information** | Guidelines recommended that patients should be provided with sufficient information with regards to their diagnosis and treatment plan, hence allowing patients themselves to better understand their behaviour and emotions and to make informed choices  *For some patients, it may be of value to as well as going through criteria texts, as the patient probably feels again in the description. The clinician can then refer to such things as the patient himself told, which provides a link between the diagnosis and the patient's reality.*  Swedish Psychiatric Association (Sweden)  *Before offering psychological treatment and, in any case, also for all other offers of the Service (drug therapies, job grants ... etc), for PGD or for the conditions of co-morbidities, clear information must be provided through a written text with the characteristics of the treatment offered.*  Mental Health Department (Italy) | 24 |
| **Hope** | Clinician’s responses and treatment options were recommended by some guidelines to reflect optimism and hope, helping to keep patients engaged in treatment  *When working with people with Severe Personality Disorder, you need to address the different treatment options in an atmosphere of trust and optimism, explaining that healing is possible and can be achieved*  Mental Health Department (Italy)  *Users felt that there needs to be acknowledgement by professionals that personality disorder is treatable: a negative experience on initial referral to a psychiatrist makes engagement less likely.*  PD: No longer a diagnosis of exclusion (UK) | 11 |
| *Helps to speak about feelings* | Furthermore, some guidelines also discussed the act of verbalising feelings being cathartic  *The ability to reﬂect and discuss in a reﬂective and differentiated way about thoughts, feelings and relationships has been operationalized with the concept of function reﬂexive. It has been recognized that an improvement in mentalization capacity is a central component of efﬁcacy in BPD psychotherapy*  SAPP: Treatment recommendations for BPD(Switzerland) | 7 |
| **Evaluation of treatment** | Apart from providing treatment, some guidelines also noted the importance of evaluating treatment administered at various timepoints  *Therapeutic success is determined primarily by a reduction in the predominant symptomatology as well as an improvement in the living situation psychosocial. These areas are regularly assessed together by the therapist and the patient.*  SAPP: Treatment recommendations for BPD(Switzerland)  *Monitor the effects of treatment on a broad range of outcomes including personal, social and occupational functioning, drug and alcohol use, self-harm, depression and the symptoms of personality disorder*  Illawarra Health and Medical Research Institute (Australia) | 10 |
| **Stepped care or clinical staging** | Some of the guidelines recommended adopting a stepped care approached in provision of treatment, hence ensuring that limited resources are used appropriately, and severity of symptoms are met with equal care  *Stepped care’ approaches have been advocated to overcome the problem of poor access to psychological therapy services due to the limited availability of trained therapists.*  National Health and Medical Research Council (Australia)  *When a person is referred, the stepped care approach is used to match their needs with the right level of support; the individual only ‘steps up’ to intensive / specialist services as their needs require.*  HSC: Regional Care Pathway for PD (Ireland)  *The staged care models are based on a population-based approach covering all the health needs of a population, from prevention to specialized treatment. These needs are assessed taking into account the complexity and intensity of the symptoms, the personal and social characteristics and user preferences. Models of care by step are most often described as the interaction of two principles (Richards and al., 2012): The principle of "least burden" : a convincing (effective) intervention of low intensity is offered ﬁrst and higher intensity treatments are offered to individuals at risk to themselves or to others, who have a history of treatment failure or that do not show improvement following the initial intervention.*  Centre national d'excellence en santé mentale (Canada) | 17 |
| **Well Structured and clear interventions** | Some guidelines also recommended that interventions should have a clear structure, ensuring that the treatment is administered as intended, and to providing a timeline of the treatment  *severely personality disordered individuals derive from their treatment comes through their experience of being involved in a well-constructed, well-structured and coherent interpersonal endeavour.*  PD: No longer a diagnosis of exclusion (UK)  *Services should ensure that all staff providing psychosocial or pharmacological interventions for the treatment or prevention of antisocial personality disorder are competent and properly qualified and supervised, and that they adhere closely to the structure and duration of the interventions as set out in the relevant treatment manuals. This should be achieved through: l use of competence frameworks based on relevant treatment manuals l routine use of sessional outcome measures l routine direct monitoring and evaluation of staff adherence, for example through the use of video and audio tapes and external audit and scrutiny where appropriate.*  NICE: Anti-social PD Treatment and Management (UK)  It was also asserted that providing patients with a structure would create a sense of regularity and control over their own lives    *It is recommended that as much structure and structure as possible be maintained around patients with personality disorders to create regularity.*  TRIMBOS Institute (Netherlands) | 22 |
| *End of contact* | Having a discussion on the closure of therapy was also recommended to be addressed by a few guidelines. This would ensure that patients do not experience a sudden loss of support.  *In the event that psychotherapy comes to an end, prepare with sensitivity well in advance, consider using a symbolic ending (such as a card or letter), summarise the therapy and invite the consumer to also summarise. In the case of unplanned termination (for example, clinician changing jobs), handle with care and sensitivity.*  SANE (Australia)  *The treating clinician can help the person cope by emphasising any progress that the person has made towards recovery, clearly expressing confidence in the person’s ability to manage their life now and after the end of treatment, encouraging the person to think about future goals and challenges and how they will approach these, and supporting the person to identify other sources of support*  National Health and Medical Research Council (Australia) | 10 |
| **Holistic care and general well-being** | Some guidelines discussed the feasibility of providing support or signposting patients to support beyond clinical needs, addressing issues that are more logistical in nature  *People with BPD should be encouraged to proactively use community and voluntary sector services to help them address specific social problems that that may be impacting on their mental wellbeing including: advice agencies; addressing debt; housing problems; parenting support; physical health and wellbeing; training and employment opportunities.*  HSC: Regional Care Pathway for PD (Ireland)  *Practical support in the ﬁeld of ﬁnances, housing, household and administration may then be necessary. Hereby Social Work or a personal budget (PGB) can make a valuable contribution.*  TRIMBOS Institute (Netherlands) | 9 |
| *General Well-being* | Apart from treating clinical symptoms, some guidelines also recommended to treat other aspects of the patient’s health  *Good nutrition and physical activity are associated with emotional wellbeing*  COPE: Perinatal Mental Health Guide (Australia)  *A practitioner can point out that a healthy lifestyle, such as getting enough sleep,*  GGZ: Standaarden (Netherlands)  *To work for health promotion, where one tries to help to inﬂuence the individual's lifestyle and behavior to promote health, is an important part of nursing*  Swedish Psychiatric Association (Sweden) | 5 |
| *Skills based intervention* | Some guidelines also included recommendations of teaching functional skills to patients on top of existing treatment, equipping patients with necessary skills as they transit out of care  *Services will value the individual as a person and help them develop a positive and solution focused approach to the management of their needs. Services will work to enable the individual to maximise their personal strengths, resources and talents.*  HSC: Regional Care Pathway for PD (Ireland)  *Support for people with mental health problems can be sought to improve their ability to function, to interrupt their vocational studies, to ﬁnd work motivation and to ﬁnd a suitable one.*  FMSD: Unstable Personality (Finland) | 17 |
