## Supporting Information 1 for "Evaluation of international guidance for the community treatment of complex emotional needs: A systematic review"

**Supplementary Material 1. Complete search strategy used in each bibliographic database.**

**Original broad scope searches up to March 2019:**

| **Database** | **Hits** |
| --- | --- |
| **MEDLINE** | **1861** |
| **Embase** | **2815** |
| **PsycINFO** | **2666** |
| **CINAHL** | **435** |
| **SPP** | **254** |
| **HMIC** | **125** |
| **ASSIA** | **616** |
| **Total** | **8772** |
| **-duplicates** |  |
| **N screened** |  |

**UPDATED SEARCH: Guidance focus**

| **Database** | **Hits** |
| --- | --- |
| **MEDLINE** | **58** |
| **Embase** | **73** |
| **PsycINFO** | **33** |
| **CINAHL** | **5** |
| **SPP** | **32** |
| **HMIC** | **1** |
| **ASSIA** | **0** |
| **Total** | **202** |
| **-duplicates** | **37** |
| **N to screen** | **165** |

At title and abstract, 13 articles were included. (Initial screening by 2 reviewers) 30^th^ April 2021.

**MEDLINE**

Database: Ovid MEDLINE(R) and Epub Ahead of Print, In-Process & Other Non-Indexed Citations and Daily <2019 to April 20,

2021>

Search Strategy:

--------------------------------------------------------------------------------

1     exp *Personality Disorders/ (29366)

2     ((personality or character*) adj3 disorder$).ti,ab,kw. (71430)

3     "axis II".ti,ab,kw. (2004)

4     ("Complex trauma" or CPTSD or "complex post-traumatic stress disorder").ti,ab,kw. (676)

5     (Complex adj (needs or mental)).ti,ab,kw. (2294)

6     *Self-Injurious Behavior/ (6106)

7     (Self-harm or self-injury).ti,ab,kw. (8861)

8     (emotion* adj2 (regulation or dysregulation or unstable or instability)).ti,ab,kw. (12940)

9     1 or 2 or 3 or 4 or 5 or 6 or 7 or 8 (114182)

10     Community Health Services/ (32148)

11     Community Mental Health Services/ (18725)

12     ((commun$ adj5 (mental health or model$1 or pathway$1 or program$ or evaluat$ or intervention$ or implement$)) or

camhs or cmht$1).ti,ab,kw. (90036)

13     (community adj5 (agenc$ or care or center$ or centre$ or clinic$ or consultant$ or doctor$ or employee$ or

expert$ or facilitator$ or healthcare or instructor$ or leader$ or manager$ or mentor$ or nurs$ or personnel$ or

pharmacy or pharmacist$ or psychiatrist$ or psychologist$ or psychotherapist$ or specialist$ or skill$ or staff$ or

team$ or therapist$ or tutor$ or visit$ or worker$ or group$ or independent or (peer$ adj3 support$) or survivor or

outpatient$ or "out patient$")).ti,ab,kw. (108218)

14     (commun$ adj5 (service or hub$ or based or deliver$ or interact$ or led or maintenance or mediat$ or operated or

provides or provider$ or run or setting$ or support or rehab$ or therap$ or service$ or treatment or management or

assessment or assistance or care or day or week)).ti,ab,kw. (232294)

15     (Independent sector or ((non institutional$ or noninstitution$) adj2 (sector$ or setting$))).ti,ab,kw. (385)

16     (network or outreach or ((specialist or day or whole) adj3 service)).ti,ab,kw. (413944)

17     10 or 11 or 12 or 13 or 14 or 15 or 16 (740758)

18     exp clinical pathway/ (7094)

19     exp clinical protocol/ (173868)

20     exp consensus/ (14870)

21     exp consensus development conference/ (12274)

22     exp consensus development conferences as topic/ (2920)

23     critical pathways/ (7094)

24     exp guideline/ (35605)

25     guidelines as topic/ (40988)

26     exp practice guideline/ (28533)

27     health planning guidelines/ (4130)

28     (guideline or practice guideline or consensus development conference or consensus development conference,

NIH).pt. (45200)

29     (position statement* or policy statement* or practice parameter* or best practice*).ti,ab,kf,kw. (36700)

30     (standards or guideline or guidelines).ti,kf,kw. (117339)

31     ((practice or treatment* or clinical) adj guideline*).ab. (43391)

32     (CPG or CPGs).ti. (5946)

33     consensus*.ti,kf,kw. (28592)

34     consensus*.ab. /freq=2 (27584)

35     ((critical or clinical or practice) adj2 (path or paths or pathway or pathways or protocol*)).ti,ab,kf,kw.

(21953)

36     recommendat*.ti,kf,kw. (44852)

37     (care adj2 (standard or path or paths or pathway or pathways or map or maps or plan or plans)).ti,ab,kf,kw.

(65286)

38     18 or 19 or 20 or 21 or 22 or 23 or 24 or 25 or 26 or 27 or 28 or 29 or 30 or 31 or 32 or 33 or 34 or 35 or 36 or 37 (566450)

39	9 and 17 and 38 (207)

40     limit 39 to dt="20190301 -20210420" (58)

**EMBASE**

Database(s): **Embase** 2019 to 2021 April  
Search Strategy:

| **#** | **Searches** | **Results** |
| --- | --- | --- |
| 1 | exp *personality disorder/ | 27910 |
| 2 | ((personality or character*) adj3 disorder$).ti,ab,kw. | 103626 |
| 3 | "axis II".ti,ab,kw. | 2687 |
| 4 | ("Complex trauma" or CPTSD or "complex post-traumatic stress disorder").ti,ab,kw. | 816 |
| 5 | (Complex adj (needs or mental)).ti,ab,kw. | 3257 |
| 6 | *automutilation/ | 8193 |
| 7 | (Self-harm or self-injury).ti,ab,kw. | 11828 |
| 8 | (emotion* adj2 (regulation or dysregulation or unstable or instability)).ti,ab,kw. | 118124 |
| 9 | 1 or 2 or 3 or 4 or 5 or 6 or 7 or 8 | 150994 |
| 10 | *community care/ | 19473 |
| 11 | *mental health service/ | 26320 |
| 12 | ((commun$ adj5 (mental health or model$1 or pathway$1 or program$ or evaluat$ or intervention$ or implement$)) or camhs or cmht$1).ti,ab,kw. | 112990 |
| 13 | (commun$ adj5 (service or hub$ or based or deliver$ or interact$ or led or maintenance or mediat$ or operated or provides or provider$ or run or setting$ or support or rehab$ or therap$ or service$ or treatment or management or assessment or assistance or care or day or week)).ti,ab,kw. | 303781 |
| 14 | (community adj5 (agenc$ or care or center$ or centre$ or clinic$ or consultant$ or doctor$ or employee$ or expert$ or facilitator$ or healthcare or instructor$ or leader$ or manager$ or mentor$ or nurs$ or personnel$ or pharmacy or pharmacist$ or psychiatrist$ or psychologist$ or psychotherapist$ or specialist$ or skill$ or staff$ or team$ or therapist$ or tutor$ or visit$ or worker$ or group$ or independent or (peer$ adj3 support$) or survivor or outpatient$ or "out patient$")).ti,ab,kw. | 147259 |
| 15 | (Independent sector or ((non institutional$ or noninstitution$) adj2 (sector$ or setting$))).ti,ab,kw. | 425 |
| 16 | (network or outreach or ((specialist or day or whole) adj3 service)).ti,ab,kw. | 529990 |
| 17 | 10 or 11 or 12 or 13 or 14 or 15 or 16 | 944325 |
| 18 | exp clinical pathway/ | 8618 |
| 19 | exp clinical protocol/ | 103333 |
| 20 | exp consensus/ | 81305 |
| 21 | exp consensus development conference/ | 23877 |
| 22 | exp consensus development conferences as topic/ | 23877 |
| 23 | critical pathways/ | 8618 |
| 24 | exp practice guideline/ | 590999 |
| 25 | health planning guidelines/ | 90528 |
| 26 | (guideline or practice guideline or consensus development conference or consensus development conference, NIH).pt. | 0 |
| 27 | (position statement* or policy statement* or practice parameter* or best practice*).ti,ab,kw. | 53641 |
| 28 | (standards or guideline or guidelines).ti,kw. | 160505 |
| 29 | ((practice or treatment* or clinical) adj guideline*).ab. | 66667 |
| 30 | (CPG or CPGs).ti. | 7188 |
| 31 | consensus*.ti,kw. | 36127 |
| 32 | consensus*.ab. /freq=2 | 36910 |
| 33 | ((critical or clinical or practice) adj2 (path or paths or pathway or pathways or protocol*)).ti,ab,kw. | 34196 |
| 34 | recommendat*.ti,kw. | 55988 |
| 35 | (care adj2 (standard or path or paths or pathway or pathways or map or maps or plan or plans)).ti,ab,kw. | 115667 |
| 36 | 18 or 19 or 20 or 21 or 22 or 23 or 24 or 25 or 26 or 27 or 28 or 29 or 30 or 31 or 32 or 33 or 34 or 35 | 1057179 |
| 37 | 9 and 17 and 36 | 560 |
| 38 | limit 37 to dd="20190301 -20210420" | 73 |

**HMIC**

Database(s): **HMIC Health Management Information Consortium** 2019 to 2021 April 
Search Strategy:

| **#** | **Searches** | **Results** |
| --- | --- | --- |
| 1 | exp *Personality Disorders/ | 0 |
| 2 | ((personality or character*) adj3 disorder$).ti,ab,hw. | 569 |
| 3 | "axis II".ti,ab,hw. | 12 |
| 4 | ("Complex trauma" or CPTSD or "complex post-traumatic stress disorder").ti,ab,hw. | 2 |
| 5 | (Complex adj (needs or mental)).ti,ab,hw. | 446 |
| 6 | *Self-Injurious Behavior/ | 0 |
| 7 | (Self-harm or self-injury).ti,ab,hw. | 701 |
| 8 | (emotion* adj2 (regulation or dysregulation or unstable or instability)).ti,ab,hw. | 357 |
| 9 | 1 or 2 or 3 or 4 or 5 or 6 or 7 or 8 | 2027 |
| 10 | Community Health Services/ | 1763 |
| 11 | Community Mental Health Services/ | 971 |
| 12 | ((commun$ adj5 (mental health or model$1 or pathway$1 or program$ or evaluat$ or intervention$ or implement$)) or camhs or cmht$1).ti,ab,hw. | 7496 |
| 13 | (community adj5 (agenc$ or care or center$ or centre$ or clinic$ or consultant$ or doctor$ or employee$ or expert$ or facilitator$ or healthcare or instructor$ or leader$ or manager$ or mentor$ or nurs$ or personnel$ or pharmacy or pharmacist$ or psychiatrist$ or psychologist$ or psychotherapist$ or specialist$ or skill$ or staff$ or team$ or therapist$ or tutor$ or visit$ or worker$ or group$ or independent or (peer$ adj3 support$) or survivor or outpatient$ or "out patient$")).ti,ab,hw. | 23555 |
| 14 | (commun$ adj5 (service or hub$ or based or deliver$ or interact$ or led or maintenance or mediat$ or operated or provides or provider$ or run or setting$ or support or rehab$ or therap$ or service$ or treatment or management or assessment or assistance or care or day or week)).ti,ab,hw. | 28610 |
| 15 | (Independent sector or ((non institutional$ or noninstitution$) adj2 (sector$ or setting$))).ti,ab,hw. | 977 |
| 16 | (network or outreach or ((specialist or day or whole) adj3 service)).ti,ab,hw. | 5135 |
| 17 | 10 or 11 or 12 or 13 or 14 or 15 or 16 | 39831 |
| 18 | exp clinical pathway/ | 1257 |
| 19 | exp clinical protocol/ | 62 |
| 20 | [exp consensus/] | 0 |
| 21 | exp consensus development conference/ | 29 |
| 22 | [exp consensus development conferences as topic] | 0 |
| 23 | critical pathways/ | 0 |
| 24 | exp guideline/ | 7169 |
| 25 | guidelines as topic/ | 0 |
| 26 | exp practice guideline/ | 1512 |
| 27 | health planning guidelines/ | 0 |
| 28 | (guideline or practice guideline or consensus development conference or consensus development conference, NIH).pt. | 0 |
| 29 | (position statement* or policy statement* or practice parameter* or best practice*).ti,ab,hw. | 2205 |
| 30 | (standards or guideline or guidelines).ti,hw. | 15355 |
| 31 | ((practice or treatment* or clinical) adj guideline*).ab. | 1393 |
| 32 | (CPG or CPGs).ti. | 1 |
| 33 | consensus*.ti,hw. | 399 |
| 34 | consensus*.ab. /freq=2 | 331 |
| 35 | ((critical or clinical or practice) adj2 (path or paths or pathway or pathways or protocol*)).ti,ab,hw. | 443 |
| 36 | recommendat*.ti,hw. | 1292 |
| 37 | (care adj2 (standard or path or paths or pathway or pathways or map or maps or plan or plans)).ti,ab,hw. | 3817 |
| 38 | 18 or 19 or 20 or 21 or 22 or 23 or 24 or 25 or 26 or 27 or 28 or 29 or 30 or 31 or 32 or 33 or 34 or 35 or 36 or 37 | 23110 |
| 39 | 9 and 17 and 38 | 31 |
| 40 | limit 39 to yr="2019 -Current" | 1 |

**SPP**

Database(s): **Social Policy and Practice** 20210413 
Search Strategy:

| **#** | **Searches** | **Results** |
| --- | --- | --- |
| 1 | [exp *Personality Disorders/] | 0 |
| 2 | ((personality or character*) adj3 disorder$).ti,ab,hw. | 1342 |
| 3 | "axis II".ti,ab,hw. | 22 |
| 4 | ("Complex trauma" or CPTSD or "complex post-traumatic stress disorder").ti,ab,hw. | 85 |
| 5 | (Complex adj (needs or mental)).ti,ab,hw. | 1599 |
| 6 | [*Self-Injurious Behavior/] | 0 |
| 7 | (Self-harm or self-injury).ti,ab,hw. | 1656 |
| 8 | (emotion* adj2 (regulation or dysregulation or unstable or instability)).ti,ab,hw. | 351 |
| 9 | 1 or 2 or 3 or 4 or 5 or 6 or 7 or 8 | 4862 |
| 10 | [Community Health Services/] | 0 |
| 11 | [Community Mental Health Services/] | 0 |
| 12 | ((commun$ adj5 (mental health or model$1 or pathway$1 or program$ or evaluat$ or intervention$ or implement$)) or camhs or cmht$1).ti,ab,hw. | 9830 |
| 13 | (community adj5 (agenc$ or care or center$ or centre$ or clinic$ or consultant$ or doctor$ or employee$ or expert$ or facilitator$ or healthcare or instructor$ or leader$ or manager$ or mentor$ or nurs$ or personnel$ or pharmacy or pharmacist$ or psychiatrist$ or psychologist$ or psychotherapist$ or specialist$ or skill$ or staff$ or team$ or therapist$ or tutor$ or visit$ or worker$ or group$ or independent or (peer$ adj3 support$) or survivor or outpatient$ or "out patient$")).ti,ab,hw. | 26373 |
| 14 | (commun$ adj5 (service or hub$ or based or deliver$ or interact$ or led or maintenance or mediat$ or operated or provides or provider$ or run or setting$ or support or rehab$ or therap$ or service$ or treatment or management or assessment or assistance or care or day or week)).ti,ab,hw. | 36494 |
| 15 | (Independent sector or ((non institutional$ or noninstitution$) adj2 (sector$ or setting$))).ti,ab,hw. | 583 |
| 16 | (network or outreach or ((specialist or day or whole) adj3 service)).ti,ab,hw. | 7398 |
| 17 | 10 or 11 or 12 or 13 or 14 or 15 or 16 | 49575 |
| 18 | [exp clinical pathway/] | 0 |
| 19 | [exp clinical protocol/] | 0 |
| 20 | [exp consensus/] | 0 |
| 21 | [exp consensus development conference/] | 0 |
| 22 | [exp consensus development conferences as topic/] | 0 |
| 23 | [critical pathways/] | 0 |
| 24 | [exp guideline/] | 0 |
| 25 | [guidelines as topic/] | 0 |
| 26 | [exp practice guideline/] | 0 |
| 27 | [health planning guidelines/] | 0 |
| 28 | (guideline or practice guideline or consensus development conference or consensus development conference, NIH).pt. | 0 |
| 29 | (position statement* or policy statement* or practice parameter* or best practice*).ti,ab,hw. | 3914 |
| 30 | (standards or guideline or guidelines).ti,hw. | 10595 |
| 31 | ((practice or treatment* or clinical) adj guideline*).ab. | 604 |
| 32 | (CPG or CPGs).ti. | 0 |
| 33 | consensus*.ti,hw. | 171 |
| 34 | consensus*.ab. /freq=2 | 136 |
| 35 | ((critical or clinical or practice) adj2 (path or paths or pathway or pathways or protocol*)).ti,ab,hw. | 93 |
| 36 | recommendat*.ti,hw. | 920 |
| 37 | (care adj2 (standard or path or paths or pathway or pathways or map or maps or plan or plans)).ti,ab,hw. | 2647 |
| 38 | 18 or 19 or 20 or 21 or 22 or 23 or 24 or 25 or 26 or 27 or 28 or 29 or 30 or 31 or 32 or 33 or 34 or 35 or 36 or 37 | 18068 |
| 39 | 9 and 17 and 38 | 64 |
| 40 | limit 39 to up="201903 -202104" | 32 |

**PsycINFO**

Database(s): **PsycINFO** March Week 1 2019 to 2021 April
Search Strategy:

| **#** | **Searches** | **Results** |
| --- | --- | --- |
| 1 | exp *Personality Disorders/ | 24721 |
| 2 | ((personality or character*) adj3 disorder$).ti,ab. | 46126 |
| 3 | "axis II".ti,ab. | 2706 |
| 4 | ("Complex trauma" or CPTSD or "complex post-traumatic stress disorder").ti,ab. | 1022 |
| 5 | (Complex adj (needs or mental)).ti,ab. | 2126 |
| 6 | *Self-Injurious Behavior/ | 3834 |
| 7 | (Self-harm or self-injury).ti,ab. | 9057 |
| 8 | (emotion* adj2 (regulation or dysregulation or unstable or instability)).ti,ab. | 19653 |
| 9 | 1 or 2 or 3 or 4 or 5 or 6 or 7 or 8 | 82510 |
| 10 | *Community Mental Health Services/ | 6722 |
| 11 | *Community Services/ | 13964 |
| 12 | ((commun$ adj5 (mental health or model$1 or pathway$1 or program$ or evaluat$ or intervention$ or implement$)) or camhs or cmht$1).ti,ab. | 63335 |
| 13 | (community adj5 (agenc$ or care or center$ or centre$ or clinic$ or consultant$ or doctor$ or employee$ or expert$ or facilitator$ or healthcare or instructor$ or leader$ or manager$ or mentor$ or nurs$ or personnel$ or pharmacy or pharmacist$ or psychiatrist$ or psychologist$ or psychotherapist$ or specialist$ or skill$ or staff$ or team$ or therapist$ or tutor$ or visit$ or worker$ or group$ or independent or (peer$ adj3 support$) or survivor or outpatient$ or "out patient$")).ti,ab. | 59070 |
| 14 | (commun$ adj5 (service or hub$ or based or deliver$ or interact$ or led or maintenance or mediat$ or operated or provides or provider$ or run or setting$ or support or rehab$ or therap$ or service$ or treatment or management or assessment or assistance or care or day or week)).ti,ab. | 131667 |
| 15 | (Independent sector or ((non institutional$ or noninstitution$) adj2 (sector$ or setting$))).ti,ab. | 220 |
| 16 | (network or outreach or ((specialist or day or whole) adj3 service)).ti,ab. | 102881 |
| 17 | 10 or 11 or 12 or 13 or 14 or 15 or 16 | 283974 |
| 18 | [exp clinical pathway/] | 0 |
| 19 | [exp clinical protocol/] | 0 |
| 20 | [exp consensus/] | 0 |
| 21 | [exp consensus development conference/] | 0 |
| 22 | [exp consensus development conferences as topic/] | 0 |
| 23 | critical pathways/ | 0 |
| 24 | [exp guideline/] | 0 |
| 25 | guidelines as topic/ | 0 |
| 26 | [exp practice guideline/] | 0 |
| 27 | health planning guidelines/ | 0 |
| 28 | (guideline or practice guideline or consensus development conference or consensus development conference, NIH).pt. | 0 |
| 29 | (position statement* or policy statement* or practice parameter* or best practice*).ti,ab,hw,id. | 19312 |
| 30 | (standards or guideline or guidelines).ti,hw,id. | 31701 |
| 31 | ((practice or treatment* or clinical) adj guideline*).ab. | 7996 |
| 32 | (CPG or CPGs).ti. | 136 |
| 33 | [consensus*.ti,hw.id.] | 0 |
| 34 | consensus*.ab. /freq=2 | 5024 |
| 35 | ((critical or clinical or practice) adj2 (path or paths or pathway or pathways or protocol*)).ti,ab,hw,id. | 2152 |
| 36 | recommendat*.ti,hw,id. | 9894 |
| 37 | (care adj2 (standard or path or paths or pathway or pathways or map or maps or plan or plans)).ti,ab,hw,id. | 9042 |
| 38 | 18 or 19 or 20 or 21 or 22 or 23 or 24 or 25 or 26 or 27 or 28 or 29 or 30 or 31 or 32 or 33 or 34 or 35 or 36 or 37 | 77597 |
| 39 | 9 and 17 and 38 | 157 |
| 40 | limit 39 to up="20190301 -20210420" | 33 |

**ASSIA**

**((MAINSUBJECT.EXACT("Personality disorders") OR (ti(((personality OR character*) NEAR/2 disorder*)) OR ab(((personality OR character*) NEAR/2 disorder*))) OR (ti("axis II") OR ab("axis II")) OR (ti(("Complex trauma" OR CPTSD OR "complex post-traumatic stress disorder")) OR ab(("Complex trauma" OR CPTSD OR "complex post-traumatic stress disorder"))) OR (ti((Complex NEAR/1 (needs OR mental))) OR ab((Complex NEAR/1 (needs OR mental)))) OR (ti((Self-harm OR self-injury)) OR ab((Self-harm OR self-injury))) OR (ti((emotion* NEAR/1 (regulation OR dysregulation OR unstable OR instability))) OR ab((emotion* NEAR/1 (regulation OR dysregulation OR unstable OR instability))))) AND (MAINSUBJECT.EXACT("Community health services") OR MAINSUBJECT.EXACT("Community mental health services") OR (ti(commun*) OR ab(commun*)) OR (ti((Independent sector)) OR ab((Independent sector))) OR (ti((non institutional* OR noninstitution*)) OR ab((non institutional* OR noninstitution*)))) AND (ti((clinical NEAR/1 (guideline OR pathway))) OR ab((clinical NEAR/1 (guideline OR pathway)))))) AND pd(20190101-2021xxxx)**

**OR as separate searches:**

**Search Strategy**

**Set#: S1**

**Searched for: MAINSUBJECT.EXACT("Personality disorders")**

**Databases: Applied Social Sciences Index & Abstracts (ASSIA)**

**Results: 4429**

**Set#: S2**

**Searched for: ti(((personality or character*) near/2 disorder*)) OR ab(((personality or character*) near/2 disorder*))**

**Databases: Applied Social Sciences Index & Abstracts (ASSIA)**

**Results: 7813**

**Set#: S3**

**Searched for: ti(((DSM and "axis II") or "cluster B")) OR ab(((DSM and "axis II") or "cluster B"))**

**Databases: Applied Social Sciences Index & Abstracts (ASSIA)**

**Results: 452**

**Set#: S4**

**Searched for: ti(("Complex trauma" or CPTSD or "complex post-traumatic stress disorder")) OR ab(("Complex trauma" or CPTSD or "complex post-traumatic stress disorder"))**

**Databases: Applied Social Sciences Index & Abstracts (ASSIA)**

**Results:159**

**Set#: S5**

**Searched for: ti((Complex near/1 (needs or mental))) OR ab((Complex near/1 (needs or mental)))**

**Databases: Applied Social Sciences Index & Abstracts (ASSIA)**

**Results: 1595**

**Set#: S6**

**Searched for: ti((Self-harm or self-injury)) OR ab((Self-harm or self-injury))**

**Databases: Applied Social Sciences Index & Abstracts (ASSIA)**

**Results: 3100**

**Set#: S7**

**Searched for: ti((emotion* NEAR/1 (regulation or dysregulation or unstable or instability))) OR ab((emotion* NEAR/1 (regulation or dysregulation or unstable or instability)))**

**Databases: Applied Social Sciences Index & Abstracts (ASSIA)**

**Results: 4467**

**Set#: S8**

**Searched for: s1 or s2 or s3 or s4 or s5 or s6 or s7**

**Databases: Applied Social Sciences Index & Abstracts (ASSIA)**

**These databases are searched for part of your query.**

**Results: 17376**

**Set#: S9**

**Searched for: MAINSUBJECT.EXACT("Community health services")**

**Databases: Applied Social Sciences Index & Abstracts (ASSIA)**

**Results: 762**

**Set#: S10**

**Searched for: MAINSUBJECT.EXACT("Community mental health services")**

**Databases: Applied Social Sciences Index & Abstracts (ASSIA)**

**Results: 3659**

**Set#: S11**

**Searched for: ti(commun***

**) OR ab(commun* )**

**Databases: Applied Social Sciences Index & Abstracts (ASSIA)**

**Results: 123064**

**Set#: S12**

**Searched for: ti((Independent sector)) OR ab((Independent sector) )**

**Databases: Applied Social Sciences Index & Abstracts (ASSIA)**

**Results: 816**

**Set#: S13**

**Searched for: ti((non institutional* OR noninstitution*)) OR ab((non institutional* OR noninstitution*))**

**Databases: Applied Social Sciences Index & Abstracts (ASSIA)**

**Results: 1475**

**Set#: S14**

**Searched for: s9 or s10 or s11 or s13**

**Databases: Applied Social Sciences Index & Abstracts (ASSIA)**

**These databases are searched for part of your query.**

**Results: 125590**

**Set#: S15**

**Searched for: ti((clinical near/1 (guideline or pathway))) OR ab((clinical near/1 (guideline or pathway)))**

**Databases: Applied Social Sciences Index & Abstracts (ASSIA)**

**Results: 2440**

**Set#: S16**

**Searched for: s8 and s14 or s15**

**Databases: Applied Social Sciences Index & Abstracts (ASSIA)**

**These databases are searched for part of your query.**

**Results: 4445**

**Set#: S17**

**Searched for: s8 and s14 and s15**

**Databases: Applied Social Sciences Index & Abstracts (ASSIA)**

**These databases are searched for part of your query.**

**Results: 11**

**Set#: S18**

**Searched for: (s8 and s14 and s15) AND pd(20190301-20210420)**

**Databases: Applied Social Sciences Index & Abstracts (ASSIA)**

**These databases are searched for part of your query.**

**Results: 0**

**----------------------------------------------------------**

**CINAHL PLUS via EBSCO**

| **#** | **Query** | **Limiters/Expanders** | **Last Run Via** | **Results** |
| --- | --- | --- | --- | --- |
| S21 | S8 AND S15 AND S18 | Limiters - Publication Year: 2019-2021; Exclude MEDLINE records  Search modes - Boolean/Phrase | Interface - EBSCOhost Research Databases  Search Screen - Advanced Search  Database - CINAHL Plus | 5 |
| S20 | S8 AND S15 AND S18 | Limiters - Publication Year: 2019-2021  Search modes - Boolean/Phrase | Interface - EBSCOhost Research Databases  Search Screen - Advanced Search  Database - CINAHL Plus | 7 |
| S19 | S8 AND S15 AND S18 | Search modes - Boolean/Phrase | Interface - EBSCOhost Research Databases  Search Screen - Advanced Search  Database - CINAHL Plus | 24 |
| S18 | S16 OR S17 | Search modes - Boolean/Phrase | Interface - EBSCOhost Research Databases  Search Screen - Advanced Search  Database - CINAHL Plus | 44043 |
| S17 | (MM "Practice Guidelines") | Search modes - Boolean/Phrase | Interface - EBSCOhost Research Databases  Search Screen - Advanced Search  Database - CINAHL Plus | 31204 |
| S16 | TI ( (clinical N1 (guideline OR pathway)) ) OR AB ( (clinical N1 (guideline OR pathway)) ) | Search modes - Boolean/Phrase | Interface - EBSCOhost Research Databases  Search Screen - Advanced Search  Database - CINAHL Plus | 16245 |
| S15 | S9 OR S10 OR S11 OR S12 OR S13 OR S14 | Search modes - Boolean/Phrase | Interface - EBSCOhost Research Databases  Search Screen - Advanced Search  Database - CINAHL Plus | 286798 |
| S14 | TI ( (network or outreach or ((specialist or day or whole) N3 service)) ) OR AB ( (network or outreach or ((specialist or day or whole) N3 service)) ) | Search modes - Boolean/Phrase | Interface - EBSCOhost Research Databases  Search Screen - Advanced Search  Database - CINAHL Plus | 92166 |
| S13 | TI ( (Independent sector or ((non institutional* or noninstitution*) N2 (sector* or setting*))) ) OR AB ( (Independent sector or ((non institutional* or noninstitution*) N2 (sector* or setting*))) ) | Search modes - Boolean/Phrase | Interface - EBSCOhost Research Databases  Search Screen - Advanced Search  Database - CINAHL Plus | 476 |
| S12 | TI ( (commun* N5 (service or hub* or based or deliver* or interact* or led or maintenance or mediat* or operated or provides or provider* or run or setting* or support or rehab* or therap* or service* or treatment or management or assessment or assistance or care or day or week)) ) OR AB ( (commun* N5 (service or hub* or based or deliver* or interact* or led or maintenance or mediat* or operated or provides or provider* or run or setting* or support or rehab* or therap* or service* or treatment or management or assessment or assistance or care or day or week)) ) | Search modes - Boolean/Phrase | Interface - EBSCOhost Research Databases  Search Screen - Advanced Search  Database - CINAHL Plus | 142731 |
| S11 | TI ( (community N5 (agenc* or care or center* or centre* or clinic* or consultant* or doctor* or employee* or expert* or facilitator* or healthcare or instructor* or leader* or manager* or mentor* or nurs* or personnel* or pharmacy or pharmacist* or psychiatrist* or psychologist* or psychotherapist* or specialist* or skill* or staff* or team* or therapist* or tutor* or visit* or worker* or group* or independent or (peer* N3 support*) or survivor or outpatient* or "out patient*")) ) OR AB ( (community N5 (agenc* or care or center* or centre* or clinic* or consultant* or doctor* or employee* or expert* or facilitator* or healthcare or instructor* or leader* or manager* or mentor* or nurs* or personnel* or pharmacy or pharmacist* or psychiatrist* or psychologist* or psychotherapist* or specialist* or skill* or staff* or team* or therapist* or tutor* or visit* or worker* or group* or independent or (peer* N3 support*) or survivor or outpatient* or "out patient*")) ) | Search modes - Boolean/Phrase | Interface - EBSCOhost Research Databases  Search Screen - Advanced Search  Database - CINAHL Plus | 86317 |
| S10 | TI ( ((commun* N5 (mental health or model* or pathway* or program* or evaluat* or intervention* or implement*)) or camhs or cmht*) ) OR AB ( ((commun* N5 (mental health or model* or pathway* or program* or evaluat* or intervention* or implement*)) or camhs or cmht*) ) | Search modes - Boolean/Phrase | Interface - EBSCOhost Research Databases  Search Screen - Advanced Search  Database - CINAHL Plus | 56626 |
| S9 | (MM "Community Health Services") OR (MM "Community Mental Health Services") | Search modes - Boolean/Phrase | Interface - EBSCOhost Research Databases  Search Screen - Advanced Search  Database - CINAHL Plus | 22728 |
| S8 | S1 OR S2 OR S3 OR S4 OR S5 OR S6 OR S7 | Search modes - Boolean/Phrase | Interface - EBSCOhost Research Databases  Search Screen - Advanced Search  Database - CINAHL Plus | 32938 |
| S7 | TI ( (emotion* N2 (regulation or dysregulation or unstable or instability)) ) OR AB ( (emotion* N2 (regulation or dysregulation or unstable or instability)) ) | Search modes - Boolean/Phrase | Interface - EBSCOhost Research Databases  Search Screen - Advanced Search  Database - CINAHL Plus | 5757 |
| S6 | TI ( (Self-harm or self-injury) ) OR AB ( (Self-harm or self-injury) ) | Search modes - Boolean/Phrase | Interface - EBSCOhost Research Databases  Search Screen - Advanced Search  Database - CINAHL Plus | 5335 |
| S5 | TI ( (Complex N1 (needs or mental)) ) OR AB ( (Complex N1 (needs or mental)) ) | Search modes - Boolean/Phrase | Interface - EBSCOhost Research Databases  Search Screen - Advanced Search  Database - CINAHL Plus | 3863 |
| S4 | TI ( ("Complex trauma" or CPTSD or "complex post-traumatic stress disorder") ) OR AB ( ("Complex trauma" or CPTSD or "complex post-traumatic stress disorder") ) | Search modes - Boolean/Phrase | Interface - EBSCOhost Research Databases  Search Screen - Advanced Search  Database - CINAHL Plus | 353 |
| S3 | TI "axis II" OR AB "axis II" | Search modes - Boolean/Phrase | Interface - EBSCOhost Research Databases  Search Screen - Advanced Search  Database - CINAHL Plus | 581 |
| S2 | TI ( ((personality or character*) N3 disorder*) ) OR AB ( ((personality or character*) N3 disorder*) ) | Search modes - Boolean/Phrase | Interface - EBSCOhost Research Databases  Search Screen - Advanced Search  Database - CINAHL Plus | 16904 |
| S1 | (MM "Personality Disorders") | Search modes - Boolean/Phrase | Interface - EBSCOhost Research Databases  Search Screen - Advanced Search  Database - CINAHL Plus | 3278 |

**Example search strategy from first search conducted in March 2019:**

**MEDLINE**

Database: Ovid MEDLINE(R) and Epub Ahead of Print, In-Process & Other Non-Indexed Citations and Daily <1946 to March 11,

2019>

Search Strategy:

--------------------------------------------------------------------------------

1     exp *Personality Disorders/ (27538)

2     ((personality or character*) adj3 disorder$).ti,ab,kw. (61634)

3     "axis II".ti,ab,kw. (1928)

4     ("Complex trauma" or CPTSD or "complex post-traumatic stress disorder").ti,ab,kw. (479)

5     (Complex adj (needs or mental)).ti,ab,kw. (1750)

6     *Self-Injurious Behavior/ (5071)

7     (Self-harm or self-injury).ti,ab,kw. (6894)

8     (emotion* adj2 (regulation or dysregulation or unstable or instability)).ti,ab,kw. (9517)

9     1 or 2 or 3 or 4 or 5 or 6 or 7 or 8 (97844)

10     Community Health Services/ (30402)

11     Community Mental Health Services/ (18023)

12     ((commun$ adj5 (mental health or model$1 or pathway$1 or program$ or evaluat$ or intervention$ or implement$)) or

camhs or cmht$1).ti,ab,kw. (74763)

13     (community adj5 (agenc$ or care or center$ or centre$ or clinic$ or consultant$ or doctor$ or employee$ or

expert$ or facilitator$ or healthcare or instructor$ or leader$ or manager$ or mentor$ or nurs$ or personnel$ or

pharmacy or pharmacist$ or psychiatrist$ or psychologist$ or psychotherapist$ or specialist$ or skill$ or staff$ or

team$ or therapist$ or tutor$ or visit$ or worker$ or group$ or independent or (peer$ adj3 support$) or survivor or

outpatient$ or "out patient$")).ti,ab,kw. (91493)

14     (commun$ adj5 (service or hub$ or based or deliver$ or interact$ or led or maintenance or mediat$ or operated or

provides or provider$ or run or setting$ or support or rehab$ or therap$ or service$ or treatment or management or

assessment or assistance or care or day or week)).ti,ab,kw. (192572)

15     (Independent sector or ((non institutional$ or noninstitution$) adj2 (sector$ or setting$))).ti,ab,kw. (358)

16     (network or outreach or ((specialist or day or whole) adj3 service)).ti,ab,kw. (323918)

17     10 or 11 or 12 or 13 or 14 or 15 or 16 (600303)

18     Interview*.af. (354084)

19     Experience*.af. (975557)

20     qualitative.tw. (197399)

21     Qualitative Research/ (44289)

22     18 or 19 or 20 or 21 (1371328)

23     randomized controlled trial.pt. (477346)

24     controlled clinical trial.pt. (92950)

25     (randomized or randomised).ab. (522337)

26     placebo.ab. (195960)

27     clinical trials as topic.sh. (186198)

28     randomly.ab. (306881)

29     trial.ti. (195319)

30     23 or 24 or 25 or 26 or 27 or 28 or 29 (1234980)

31     Epidemiologic studies/ (7898)

32     exp case control studies/ (975354)

33     Case control.tw. (113862)

34     (cohort adj (study or studies)).tw. (171707)

35     Cohort analy$.tw. (6809)

36     (Follow up adj (study or studies)).tw. (46456)

37     (observational adj (study or studies)).tw. (89758)

38     Longitudinal.tw. (218261)

39     Retrospective.tw. (460601)

40     Cross sectional.tw. (302412)

41     Cross-sectional studies/ (287583)

42     31 or 32 or 33 or 34 or 35 or 36 or 37 or 38 or 39 or 40 or 41 (1896136)

43     30 or 42 (3018405)

44     "Surveys and Questionnaires"/ (419602)

45     survey$.tw. (578168)

46     exp clinical pathway/ (6182)

47     exp clinical protocol/ (157432)

48     exp consensus/ (10077)

49     exp consensus development conference/ (11278)

50     exp consensus development conferences as topic/ (2663)

51     critical pathways/ (6182)

52     exp guideline/ (31595)

53     guidelines as topic/ (37601)

54     exp practice guideline/ (24832)

55     health planning guidelines/ (4023)

56     (guideline or practice guideline or consensus development conference or consensus development conference,

NIH).pt. (40452)

57     (position statement* or policy statement* or practice parameter* or best practice*).ti,ab,kf,kw. (28428)

58     (standards or guideline or guidelines).ti,kf,kw. (99875)

59     ((practice or treatment* or clinical) adj guideline*).ab. (35200)

60     (CPG or CPGs).ti. (5405)

61     consensus*.ti,kf,kw. (23010)

62     consensus*.ab. /freq=2 (22280)

63     ((critical or clinical or practice) adj2 (path or paths or pathway or pathways or protocol*)).ti,ab,kf,kw.

(18056)

64     recommendat*.ti,kf,kw. (36830)

65     (care adj2 (standard or path or paths or pathway or pathways or map or maps or plan or plans)).ti,ab,kf,kw.

(50434)

66     (algorithm* adj2 (screening or examination or test or tested or testing or assessment* or diagnosis or diagnoses

or diagnosed or diagnosing)).ti,ab,kf,kw. (6704)

67     (algorithm* adj2 (pharmacotherap* or chemotherap* or chemotreatment* or therap* or treatment* or

intervention*)).ti,ab,kf,kw. (8648)

68     44 or 45 or 46 or 47 or 48 or 49 or 50 or 51 or 52 or 53 or 54 or 55 or 56 or 57 or 58 or 59 or 60 or 61 or 62 or

63 or 64 or 65 or 66 or 67 (1349303)

69     22 or 43 or 68 (4852420)

70     9 and 17 and 69 (2114)

71     limit 70 to yr="2003 -Current" (1861)
