## Supporting Information 2 for "Evaluation of international guidance for the community treatment of complex emotional needs: A systematic review"

**Supplementary Material 2. Full list of guideline databases and organisations included in this search.**

### Guideline organisations

Searches conducted on Friday May 7^th^ 2021.

Organisation: TRIP Database

URL: <https://www.tripdatabase.com/>

| Search terms | Items identified (n) | Taken forward for screening (n) |
| --- | --- | --- |
| (Personality Disorder*)  LIMIT applied to clinical guidelines | 78 | 0 |
| (Personality Disorder*)  LIMIT applied to clinical guidelines  Date Limit (2019-current) | 12 | 0 |

Organisation: PsychEXTRA

URL: <https://www.apa.org/pubs/databases/psycextra>

*Unable to access

Organisation: CPG Infobase

URL: <https://joulecma.ca/cpg/homepage>

| Search terms | Items identified (n) | Taken forward for screening (n) |
| --- | --- | --- |
| Personality Disorder | 0 | 0 |

Organisation: American Psychiatric Association Clinical Practice Guidelines

URL: <https://www.psychiatry.org/psychiatrists/practice/clinical-practice-guidelines>

| Search terms | Items identified (n) | Taken forward for screening (n) |
| --- | --- | --- |
| "Personality Disorder" and guideline | 0 | 0 |

Organisation: NICE guidance

URL: <https://www.nice.org.uk/guidance>

| Search terms | Items identified (n) | Taken forward for screening (n) |
| --- | --- | --- |
| Personality Disorder | 2 | 0 (both NICE guidelines have already been identified) |

Organisation: World Health Organisation guidelines

URL: <https://www.who.int/publications/guidelines/mental_health/en/>

| Search terms | Items identified (n) | Taken forward for screening (n) |
| --- | --- | --- |
| Manually searched the webpage | 0 | 0 |

Organisation: Guidelines international network

URL: <https://www.g-i-n.net/>

| Search terms | Items identified (n) | Taken forward for screening (n) |
| --- | --- | --- |
| (Personality Disorder*) | 1  Duplicates of previous search (5) | 0 |

Organisation: Spectrum personality disorder services

URL: https://www.spectrumbpd.com.au/

| Search terms | Items identified (n) | Taken forward for screening (n) |
| --- | --- | --- |
| Manually searched the webpage | 0 | 0 |

Organisation: Australian bipolar personality disorder foundation limited

URL: https://www.bpdfoundation.org.au/

| Search terms | Items identified (n) | Taken forward for screening (n) |
| --- | --- | --- |
| Manually searched the webpage | 1  Duplicates of previous search (2) | 0 |

Organisation: SANE Australia

URL: https://www.sane.org/

| Search terms | Items identified (n) | Taken forward for screening (n) |
| --- | --- | --- |
| Manually searched the webpage | 0 | 0 |

Organisation: Mind

URL: https://www.mindaustralia.org.au/

| Search terms | Items identified (n) | Taken forward for screening (n) |
| --- | --- | --- |
| Personality Disorder guidelines | 0 | 0 |

Organisation: British and Irish group for the study of Personality Disorders (BIGSPD)

URL: https://bigspd.org.uk/

| Search terms | Items identified (n) | Taken forward for screening (n) |
| --- | --- | --- |
| (Personality Disorder*) and Guidelines | 0 | 0 |

Organisation: European Society for the study of Personality Disorder

URL: https://www.esspd.eu/

| Search terms | Items identified (n) | Taken forward for screening (n) |
| --- | --- | --- |
| Manually searched the webpage | 0  Duplicates of previous search (8) | 0 |

Organisation: International Society for the study of Personality Disorder

URL: http://www.isspd.com/

| Search terms | Items identified (n) | Taken forward for screening (n) |
| --- | --- | --- |
| Manually searched the webpage | 0 | 0 |

Organisation: North American Society for the study of Personality Disorder

URL: https://www.nasspd.org/

| Search terms | Items identified (n) | Taken forward for screening (n) |
| --- | --- | --- |
| Manually searched the webpage | 0 | 0 |
