## Supporting Information 3 for "Evaluation of international guidance for the community treatment of complex emotional needs: A systematic review"

**Supplementary Material 3. Web search and search terms used in different language.**

### Web searching

Web searching was undertaken on April 30^th^ 2021. The web-searches are reported below following the guidance of Briscoe.

Google advanced searched: <https://www.google.com/advanced_search>

April 30^th^ 2021 / May 6^th^ 2021 (different language)

| Search terms | Items identified (n) | Taken forward for screening (n) |
| --- | --- | --- |
| English- "personality disorder" guidelines | Searched to a depth of ten pages - 21 | 21 |
| French- "trouble de la personnalité" "des lignes directrices" | Searched to a depth of four pages - 1 | 1 |
| German- “Persönlichkeitsstörung” “Richtlinien” | Searched to a depth of four pages - 2  Guidelines from KI (2) | 2 |
| Italian- linee guida "disturbo della personalità" | Searched to a depth of four pages - 1  Guidelines from KI (1) | 1 |
| Spanish- pautas para el trastorno de la personalidad | Searched to a depth of four pages - 3 | 3 |
| Danish- “retningslinjer” for “personlighedsforstyrrelse” | Searched to a depth of four pages - 3  Guidelines from KI (1) | 3 |
| Norwegian- retningslinjer for personlighetsforstyrrelse | Searched to a depth of four pages - 0 | 0 |
| Portuguese- “diretrizes” de “transtorno de personalidade” | Searched to a depth of four pages - 0 | 0 |
| Greek- οδηγίες διαταραχής της προσωπικότητας | Searched to a depth of four pages - 0 | 0 |

Dogpile: <https://www.dogpile.com/>

April 30^th^ 2021

| Search terms | Items identified (n) | Taken forward for screening (n) |
| --- | --- | --- |
| ((Personality Disorder*) and (guideline*)) | Searched to a depth of ten pages - 4  Guidelines that were found in Google= 9 | 4 |
