## Supporting Information 4 for "Evaluation of international guidance for the community treatment of complex emotional needs: A systematic review"

**Supplementary Material 4. Amended version of AGREE-II.**

AGREE-II was designed to evaluate the methodological rigour and transparency with which a guideline was developed. However, not all 23 items within the original tool were applicable to the literature identified in this review. An example would be item 15 (The recommendations are specific and unambiguous.), as international guidelines that were translated into English using Google translate might have translation issues, causing some phrases or sentences to hold little meaning in English. This is not the fault of the guideline, but the translation tool used, hence it would be inappropriate to evaluate international guidelines with reference to item 15. As stated in the user manual of AGREE-II, items may be skipped if deemed to be irrelevant. Therefore, a decision was made by all four reviewers (NW, PB, LSR, and JB) to omit evaluating all international guidelines based on item 15. Out of the 23 items from the framework, only 14 were used to evaluate guidelines that were in English and 13 were used to evaluate translated guidelines.

Points in red are those that have been excluded from our quality appraisal.

1. Domain 1: scope and purpose
   a. The overall objective(s) of the guideline is (are) specifically described
   b. The health question(s) covered by the guideline is (are) specifically described
   c. The population (patients, public, etc.) to whom the guideline is meant to apply is specifically described.
2. Domain 2: Stakeholder involvement
   a. The guideline development group includes individuals from all relevant professional groups
   b. The views and preferences of the target population (patients, public, etc.) have been sought.
   c. The target users of the guideline are clearly defined.
3. Domain 3: Rigour of development
   a. Systematic methods were used to search for evidence.
   b. The criteria for selecting the evidence are clearly described.
   c. The strengths and limitations of the body of evidence are clearly described.
   d. The methods for formulating the recommendations are clearly described.
   e. The health benefits, side effects, and risks have been considered in formulating the recommendations.
   f. There is an explicit link between the recommendations and the supporting evidence.
   g. The guideline has been externally reviewed by experts prior to its publication.
   h. A procedure for updating the guideline is provided.
4. Domain 4: Clarity of presentation
   a. The recommendations are specific and unambiguous.
   b. The different options for management of the condition or health issue are clearly presented.
   c. Key recommendations are easily identifiable.
5. Domain 5: applicabilit2
   a. The guideline describes facilitators and barriers to its application.
   b. The guideline provides advice and/or tools on how the recommendations can be put into practice.
   c. The potential resource implications of applying the recommendations have been considered.

d. The guideline presents monitoring and/or auditing criteria.

6. Domain 6: editorial independence
   a. The views of the funding body have not influenced the content of the guideline.
   b. Competing interests of guideline development group members have been recorded and addressed.
